# “Immune System Modulation with Oral Vancomycin in combination with Stereotactic Body Radiotherapy (SBRT) for medically inoperable Early-Stage Non-small Cell Lung Cancer”

**DOI:** 10.1101/2025.01.09.25319989

**Authors:** Steven J. Feigenberg, Francesca Costabile, Ceylan Tanes, Kyle Bittinger, Roddy O’Connor, Divyansh Agarwal, Giorgos Skoufos, Silvano Salaris, Artemis Hatzigeorgiou, Nektarios Kostopoulos, Shane Lloyd, Cole Friedes, Lisha Chen, Nikhil Yegya-Raman, Keith Cengel, William Levin, Bakir Valentić, Tyler Quarton, Alexander A. Shestov, Abigail Berman, Jeffrey Bradley, Amit Maity, Costantinos Koumenis, Edgar Ben-Josef, Andrea Facciabene

**Affiliations:** University of Pennsylvania, Department of Radiation Oncology.; Division of Gastroenterology, Hepatology, and Nutrition, Children’s Hospital of Philadelphia, Philadelphia, Pennsylvania, USA; University of Pennsylvania, Center for Cellular Therapy.; Massachusetts General Hospital, Harvard Medical School.; University of Thessaly, Thessaly, Greece; Istituto Zooprofilattico Sperimentale delle Venezie, 35020 Legnaro, Italy; University of Utah School of Medicine and Huntsman Cancer Institute.; Fox Chase Cancer Center, Department of Radiation Oncology.; University of Pennsylvania, the Ovarian Cancer Research Center.; The University of Texas at Dallas

## Abstract

We present the results of a randomized, open-label pilot study investigating the combination of oral vancomycin and stereotactic body radiotherapy (SBRT) in early-stage non-small cell lung cancer (NSCLC). Our findings highlight vancomycin’s safety, evidenced by the absence of Grade 3 or 4 adverse events, and its potential to enhance the antitumor efficacy of SBRT. The observed enhancement is linked to vancomycin’s modulation of the gut microbiota, which triggers significant metabolic changes and immune activation, thereby contributing to improved progression-free survival (PFS) and overall survival (OS). Patients received vancomycin (125 mg, four times daily for five weeks, starting one week prior to SBRT), which induced restructuring of the gut microbiome and significant changes in the gut metabolome. Key changes included reductions in short-chain fatty acids (SCFAs) and shifts in other immunomodulatory metabolites. These metabolic shifts were associated with the activation of dendritic cells and T cells, creating a pro-inflammatory environment conducive to strengthening SBRT’s antitumor efficacy. The combination of vancomycin and SBRT presents a novel, low-toxicity therapeutic approach for early-stage NSCLC, showing promising initial outcomes. While the results are encouraging, further research with larger cohorts is necessary to verify these findings and elucidate the underlying mechanisms that contribute to the observed clinical benefits.

**WHAT IS ALREADY KNOWN ON THIS TOPIC:** Radiation therapy is a primary treatment for early-stage non-small cell lung cancer and offers excellent local control in early-stage NSCLC, the challenges of regional and distant failures which occur in up to 50% of patients, lead to increased morbidity and mortality. The gut microbiome is increasingly recognized in cancer immunotherapy. RT can induce Immunogenic Cell Death, activating the immune system and promoting abscopal effect to impact untreated lesions. Our previous preclinical studies have shown that antibiotics like vancomycin can modulate these immune effects and enhance RT’s antitumor activity.

**WHAT THIS STUDY ADDS:** This clinical study corroborates our previous preclinical findings by demonstrating the safety of vancomycin and its potential to enhance the antitumor effects of RT, despite the small cohort size. These findings suggest that vancomycin could be strategically used to improve RT outcomes.

**HOW THIS STUDY MIGHT AFFECT RESEARCH, PRACTICE OR POLICY:** Our findings prompt further investigation into this combined treatment in a larger patient cohort to confirm enhanced progression-free survival and overall survival. Exploring the impact on distal recurrences and applying this strategy to more advanced patient stages could significantly influence future research directions and clinical practices. This approach may also guide policy towards integrating microbiome modulation strategies in standard cancer treatment protocols.

## Introduction

SBRT is a used in early-stage non-small cell lung cancer (NSCLC) for patients who are inoperable or decline surgery. While SBRT offers excellent local control in early-stage NSCLC, the challenges of regional and distant failures which occur in up to 50% of patients, lead to increased morbidity and mortality. Current guidelines mirror surgical data where adjuvant systemic therapy is recommended for tumors ≥4 cm. This benefit is not seen for smaller tumors as the morbidity and mortality of systemic therapy appears to outweigh the benefit. Furthermore, patients receiving SBRT are older with more comorbidities while at the same time likely have a greater burden of subclinical disease as nodes are not assessed making it imperative to develop more effective and less toxic systemic agents^1,2^.

The gastrointestinal tract, hosting the largest number of immune cells in the body, plays a crucial role in immune system regulation^3^. Recent studies emphasize the intestinal microbiota’s pivotal role in modulating immune responses, with disturbances linked to various autoimmune disorders^4,5^. Additionally, the gut microbiome’s composition significantly influences the efficacy of immunotherapies^6–8^. Notably, fecal microbiota transplantation (FMT) has shown promise in enhancing checkpoint blockade therapy’s antitumor efficacy, sparking interest in its potential across diverse cancer types^9,10^.

Building on this knowledge, our previous research explored the impact of gut microbiota perturbation on the efficacy of radiation therapy (RT)^11^. We discovered that oral vancomycin treatment alters the monocytes and dendritic cell features, enhancing antigen presentation and cytokine secretion, which supports the priming of an anti-tumor T cell response and ultimately improves RT outcomes. This combination therapy increases the expression of key genes involved in antigen presentation and Th1 effector T cell development, such as IFN-β and IFN-γ^12–14^. Additionally, vancomycin led to significant changes in the gut microbiota, including a decrease in alpha diversity, a reduction in Firmicutes, Bacteroidetes, and an increase in Proteobacteria^15,16^. This shift included a notable reduction in SCFAs-producing bacteria^15^, which are known for their overall anti-inflammatory effects^17^ and can counteract the synergistic effects of vancomycin and radiation therapy when supplemented externally^11^. These findings underscore the potential of microbiome modulation as a strategy to enhance anti-tumor immunity and improve clinical outcomes in cancer therapy. Based on these results we launched a clinical trial (NCT03546829). Here we present the final results of the 1st randomized open label clinical pilot study involving patients treated with SBRT for early-stage lung cancer with or without the administration of oral vancomycin.

## Materials and Methods

### Study Design and Objectives

This trial was designed as an open-label, randomized pilot study (NCT03546829). It was reviewed and approved by the Institutional Review Board at the University of Pennsylvania. All patients provided written consent to participate. The primary safety endpoint was uncontrolled diarrhea, as defined by CTCAE v5.0 > or = grade 3. This was evaluated during SBRT and at 1 month, 3 months 6 months and 12 months post SBRT. The additional primary objective of the trial was to determine if the addition of vancomycin to SBRT was safe and may increases Th1 cytokines profile. Secondary objectives included determination of vancomycin induced changes in the gut microbiota composition as well as SCFAs, measures of other inflammatory cytokines. Exploratory goals were to look for a signal for differences in local control, disease-free survival and acute and chronic toxicity in each arm.

### Inclusion/Exclusion Criteria

Adults aged 18 and older, with an ECOG performance status of 0 to 2 and diagnosed with early-stage non-small cell lung cancer (NSCLC) planning to receive SBRT were eligible. While a confirmatory biopsy was recommended, it was not mandatory based on multi-disciplinary discussions. All candidates underwent PET/CT scans within six weeks before SBRT to exclude nodal or distant metastases, and brain imaging was performed only if neurological symptoms were present. Patients were excluded if they had used antibiotics, antifungals, antivirals, or antiparasitics within four weeks prior to registration; had an active infection with a temperature over 100°F; used corticosteroids, methotrexate, or immunosuppressives in the four weeks before registration; underwent chemotherapy in the four weeks prior to or during radiotherapy; had a history of HIV, HBV, or HCV; suffered from uncontrolled gastrointestinal disorders, including inflammatory bowel disease or severe irritable bowel syndrome; had persistent gastrointestinal infections or conditions such as Clostridium difficile infection within two years or untreated Helicobacter pylori infection; underwent significant GI surgery (excluding cholecystectomy or appendectomy) within the past five years or any major bowel resection ever; or were currently using anti-diarrheal medications or probiotics.

### Treatment Approach

SBRT was administered per institutional guidelines for peripheral or centrally located tumors under 5 cm. Treatment was delivered as 50 Gy across 4 or 5 fractions on non-consecutive days, while other fractionation regimens were excluded. Patient setup involved 4D CT simulation with upper and lower vaclok bags for reproducibility and a compression belt for tumors in the lower lobe to reduce motion. Treatment volumes were defined using the 4D CT images to create an IGTV, with an isotropic 5 mm margin added to form the PTV. Established dose constraints from previous publications were followed.

### Vancomycin Administration

If subjects were randomized to the study drug arm, Vancomycin was administered at 125 mg 4 times daily starting 1 week prior to SBRT and continued for a total of 5 weeks. Medication compliance was assessed with the use of a Pill Diary case report form completed by the subjects and reviewed weekly with treating physician or research staff.

To reduce the risk to subjects, a stopping rule was implemented if more than 33% of subjects given oral vancomycin experience uncontrolled *diarrhea, as defined by CTCAE v5.0 > or = grade 3)*.

### Sample Size Determination

Our primary objective was to assess the impact of vancomycin on cytokine expression changes following SBRT, estimating these effects with a 95% confidence interval. The null hypothesis posits that SBRT combined with vancomycin does not increase expression of Th1 cytokines such as IFN-gamma, IFN-beta, and TNF-alpha, compared to SBRT alone. The alternative hypothesis suggests an increase in these cytokines with the combined treatment. The hypotheses were tested using a time-by-treatment interaction analyzed via regression, or difference of differences scores assessed with a t-test. With two groups of 20, using a Cohen’s d effect size of 0.8, we achieve 80% power at a 10% type I error rate. The detectable effect size reflects within-group score differences. Variability in these differences is influenced by the raw SD of cytokine levels and within-subject correlation. For 20 samples, the 90% confidence interval for the mean difference will span approximately 0.77 SD. For the full sample of 40, the 90% CI for correlation ranges from 0.20 (high correlation, rho=0.9) to 0.53 (no correlation).

#### Please refer to online Material and Methods for the following presented data

Metagenomic sequencing, Bioinformatics processing, Metabolomic analysis MS-TOF, Heatmap procedure, RT-PCR, Flow cytometry, ELISA, Total RNA seq., RNA sequencing data analysis, Pathway Analysis.

### Data Availability Statement

RNA sequencing data from PBMCs generated in this study are publicly available in the NCBI Gene Expression Omnibus (GEO) database under accession numbers GSE277984. Shotgun sequencing data from stool samples can be accessed in the NCBI Sequence Read Archive (SRA) under accession numbers PRJNA1153928. Other data generated in this study are available within the article and its supplementary data files. Further details and data are available upon reasonable request from the authors.

### Statistical Analysis

The primary and secondary analyses included all randomized patients (intention-to-treat analysis), except for the toxicity analysis and translational endpoints, which analyzed patients by receipt of vancomycin regardless of randomization (per protocol analysis). The number of patients who were not evaluable, who died or withdrew before treatment began were specified. Progression-free survival (PFS), overall survival (OS), and freedom from local failure, locoregional failure and distant metastases were estimated using the Kaplan-Meier method, and comparisons between the vancomycin and control groups were made using the log-rank test with a one-sided type I error rate of 0.10.

For the metagenomic data, community-level differences between control and treatment groups were assessed using the PERMANOVA test at each time point. Differences in alpha diversity, relative abundance of bacteria and relative abundance of KEGG orthologs between control and treatment groups were assessed using linear models. The difference between time points within a study group was assessed using linear mixed effects models with subject IDs as random effects. Multiple tests were corrected for false discovery rate using Benjamini-Hochberg method.

For quantitative PCR (qPCR), Flow Cytometry (FACS), Mass Spectrometry (MS), and other assays requiring direct comparisons between Control and Treatment groups, the Shapiro-Wilk test was initially conducted to evaluate the normality of data distribution within each group. Where data were confirmed to be normally distributed—or when normality could not be definitively assessed—a two-tailed independent samples t-test (specifically Welch’s t-test, which does not assume equal variances) was employed. For datasets that did not follow a normal distribution, the non-parametric Mann-Whitney U test was utilized as an alternative.

## Results

### “Patient Enrollment and Study Cohort Characteristics”

Between March 2019 and August 2021, seventeen patients signed consent and were treated on study. Unfortunately, we had to close the trial early before the planned accrual of 40 patients due to the impact of the COVID 19 pandemic that severely impacted this patient population’s willingness to participate in research. Nine patients were randomized to receive Vancomycin and SBRT, eight were treated with SBRT alone. One patient was taken off study (Vancomycin arm) as the treatment plan was altered to 15 fractions after the simulation revealed the tumor to be in an ultra central location. A second patient, randomized to vancomycin, withdrew from the study prior to starting vancomycin. Therefore, the 15 patients who were treated per protocol on study will be described in detail. Table 1 demonstrates the clinical features and the three-year control rate between the patients treated with SBRT alone compared with those treated with Vancomycin and SBRT.

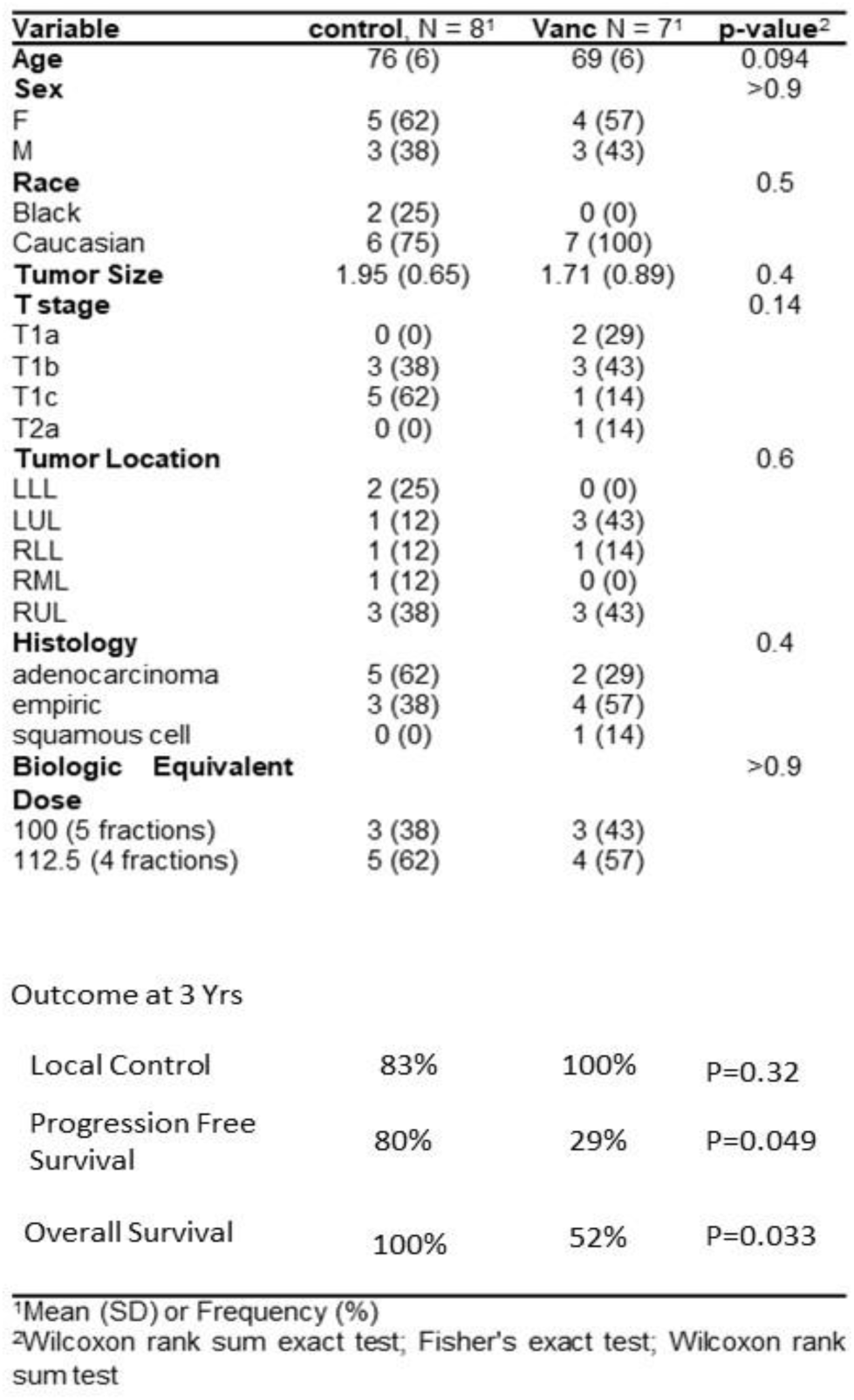

### “Tolerance and compliance of vancomycin”

Nine patients were randomized to receive Vancomycin. One patient was taken off study as the treatment plan as their tumor was ultracentral as described above. A second patient withdrew before starting vancomycin, leaving 7 patients who started Vancomycin. The most common side effects were diarrhea (4 patients), nausea (2 patients) and fatigue (1 patients). There were no grade 3 adverse events. All adverse events were grade 1 except one patient with grade 2 diarrhea and 1 patient with a grade 2 urinary tract infection. Of the 7 patients, 4 patients complete all 5 weeks of Vancomycin, while 2 patients continued treatment for 15 days and 1 continued it for 7 days. The 3 patients who did not complete the planned 5 weeks of Vancomycin stopped due to nausea in 2 patients and due to a development of a urinary tract infection in the 3^rd^ patient.

### “Oncologic Outcomes”

With a median follow up 3 years (IQR 1.7 yrs – 3.4 yrs), the 3 year local control rate, progression free survival and overall survival was 91%, 55%, and 75% respectively. Four patients have developed a recurrence of their original cancer, three of whom were treated with SBRT alone (see table 2). The most common site of failure was nodal seen in all 4 patients at 18, 20, 24 and 32 months post SBRT. One patient additionally failed locally (SBRT alone), while 3 patients additionally developed distant failure manifested by development of pleural effusion or pleural metastases simultaneously. One patient who developed a recurrence was subsequently treated with curative intent with concurrent chemoradiation therapy. Figure **1** **Left** and **1 Right** demonstrate the KM estimates for the significant improvement in progression free survival (P.F.S.) and overall survival (O.S.) from the addition of vancomycin. Further analysis of time to distant metastasis and local and locoregional failures shows a trend towards improvement; however, statistical significance was not reached, likely due to the small sample size (**Supp. Figure 1**).

**Table 2.** Details on Patients that developed a recurrence. The table provides detailed clinical information on four patients who were treated with stereotactic body radiation therapy (SBRT) for lung cancer, with some also receiving Vancomycin.

| Race | Sex | Histo | Size | Location | Dose | Vanco | Time to Failure | Site of Failure |
| --- | --- | --- | --- | --- | --- | --- | --- | --- |
| W | F | AdenoCa | 1.0 cm | RUL | 50 Gy in 4 fx | Yes | 32 months | mediastinal/ pleural studding |
| AA | F | AdenoCa | 1.1 cm | RUL | 50 Gy in 4 fx | No | 18 months | local/hilar/pleural effusion |
| W | F | Empiric | 2.1 cm | RML | 50 Gy in 5 fx | No | 20 months | hilar |
| W | F | AdenoCa | 2.5 cm | LLL | 50 Gy in 5 fx | No | 24 months | mediastinal/pleural effusion |

### “Stool and serum collection”

Blood and stool samples were collected at five designated time points throughout the study from participants in both the vancomycin group and the control group. Compliance with stool sample collection ranged from 20% to 80% in the vancomycin group, where one participant provided only one sample (20% compliance), four participants achieved 80% compliance, and two maintained 60% compliance. In the control group, three participants achieved an 80% compliance rate, two participants had a 60% compliance rate, and one participant submitted only one stool sample. Serum sample compliance varied similarly, with rates ranging from 40% to 80% across different time points. The variability in compliance can largely be attributed to the impact of the COVID-19 pandemic, which introduced significant challenges. Additionally, the predominantly older cohort found the process of self-collecting stool samples particularly burdensome, which disproportionately affected the compliance rates for stool samples.

### “Vancomycin-Induced Shifts in Microbial Diversity: A Closer Look at Antibiotic Resistance and Microbial Dynamics”

The administration of vancomycin resulted in a notable decrease in the alpha diversity of the participants’ gut microbiota which persisted over a month post SBRT time point (**Figure 2A**). Furthermore, at 3-month time point, the alpha diversity of the treatment group remained lower than the control group despite the cessation of antibiotics. The alpha diversity in the control group remained relatively stable except for a slight increase at the 3-month time point compared to baseline (**Figure 2A**). In line with previous works the use of vancomycin on patients has altered the composition of the microbiome^16^. Antibiotics had a significant effect on the microbiome, as the microbiome composition of the subjects during and after antibiotics treatment was different than the control cohort (**Supp Figure 2A**, PERMANOVA, p=0.003). Furthermore, when we investigated each time point separately, the microbiome composition at inspection time point was different between control and treatment groups (PERMANOVA, p=0.01) indicating that the effect of vancomycin persisted past the therapy window (**Figure 2B**) confirming previous works. ^15^. At the phylum level, vancomycin treatment during the SBRT induced a decrease in *Bacteroidetes* and an increase in *Proteobacteria* compared to baseline (**Figure 2C**). After vancomycin was completed, we observed a reduction in *Actinobacteria* at the inspection time point (**Figure 2C**). Various species from the Bacteroidetes phylum decreased during treatment such as B. vulgatus, B. uniformis and B. fragilis. This resulted in an increase in E. coli during the vancomycin treatment. Three species belonging to the *Lachnospiraceae* family and additional belonging *Ruminococcaceae*, which contains numerous butyrate producers were lower at the inspection time point in the treatment group compared to controls (**Figure 2D**). Akkermansia, Veillonella, Megasphaera and Enterococcus, previously identified as important in the antitumor response mediated by ICI, ACT as well GVHD for Lymphoma patients receiving allo-transplantation, only showed changes before false discovery rate correction during the vancomycin use^6,18–20^. *Veillonella* levels showed trends of higher levels compared to control and were higher during the treatment compared to baseline whereas *Akkermansia* levels showed trends of lower levels during SBRT (**Supp. Figure 2B**).

**Figure 1.**
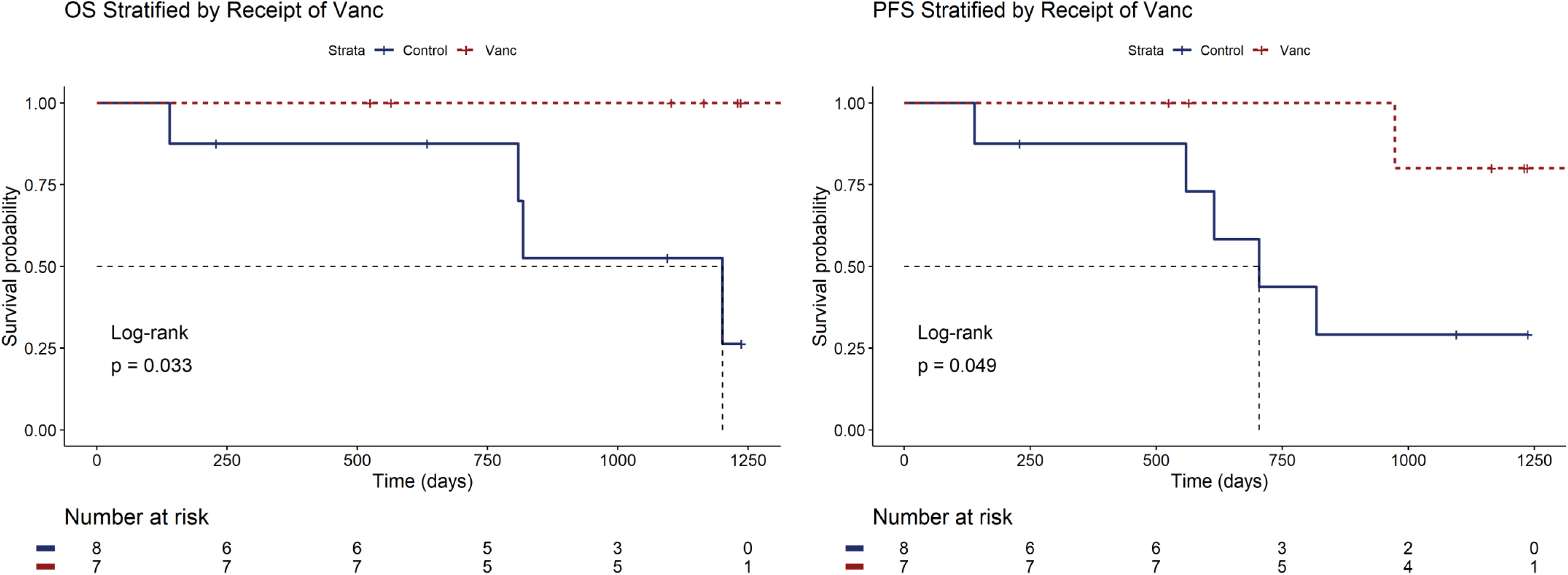
Oncologic Outcomes. **Left.** O.S. as a function of Vancomycin use. **Right.** P.F.S as a function of Vancomycin use.

**Figure 2:**
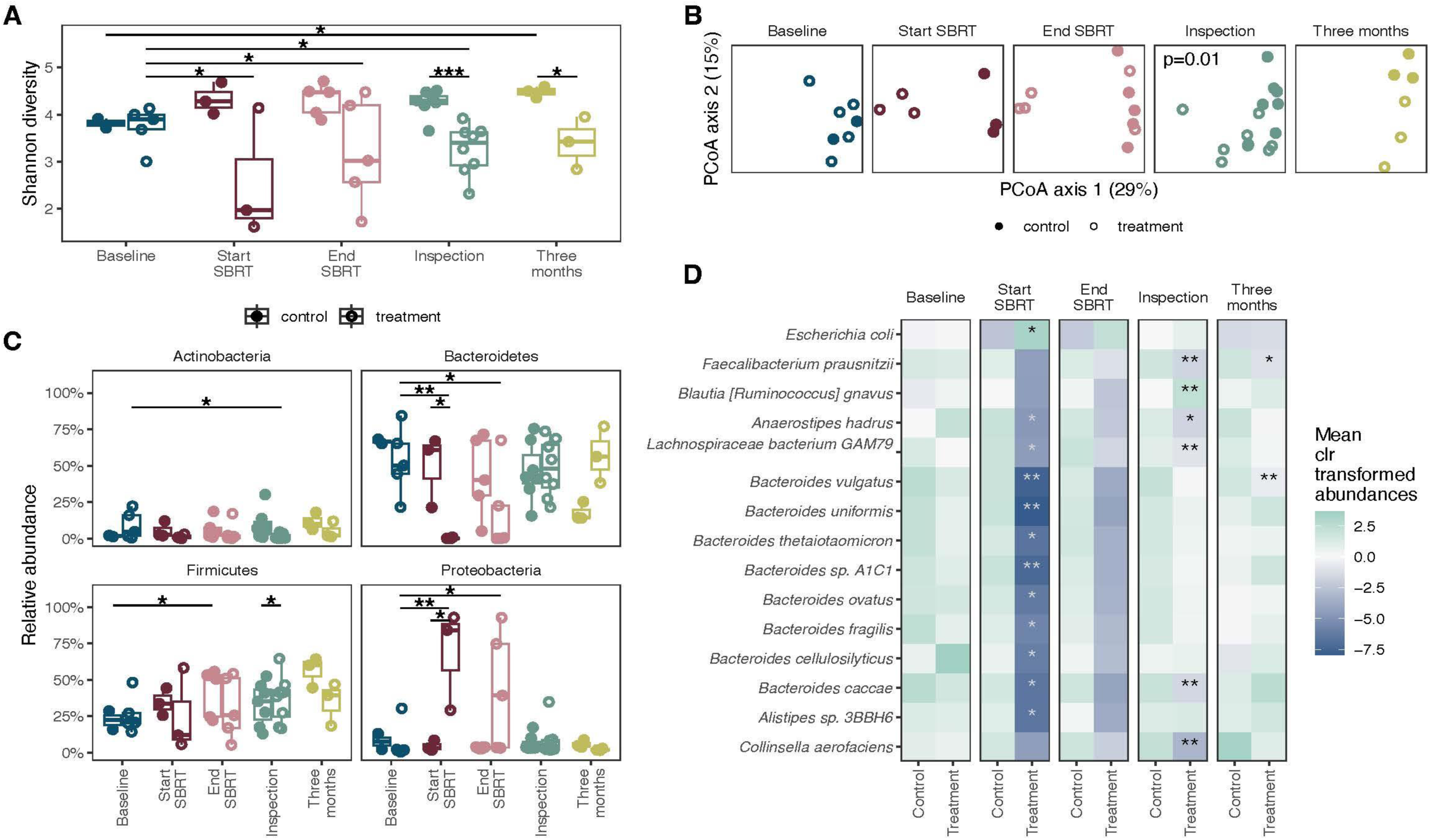
Effect of vancomycin treatment on bacterial abundances. A) Shannon diversity of samples. B) Principal coordinate analysis on Bray-Curtis distances of taxonomic abundances of bacteria at the different time points assessed. C) Relative abundance of four major phylum in the gut. Differences across time were tested using linear mixed effects models and differences between control and treatment groups were tested using linear models. ***<0.001, **<0.01, *<0.05. D) Mean of center log ratio (clr) transformed abundances at each time point. The stars mark the bacteria that are different between the control and treatment groups for that time point using linear models.

### “From Microbiome to Metabolome: Tracing the Effects of Vancomycin Treatment through KEGG Pathways Analysis”

The relative abundance of orthologs and the relevant pathways were analyzed to determine the microbial shifts on the host metabolism using the Kyoto Encyclopedia of Genes and Genomes (KEGG) database^21^. This approach enabled us to elucidate the potential impact of vancomycin-induced changes in the microbiome on the host’s metabolic landscape, with a particular focus on pathways known to affect the immune system. The gene composition of subjects during and after antibiotic treatment differed from that of the control cohort (PERMANOVA, p=0.03; **Figure 3A**). However, when analyzing individual time points, we observed a trend but did not detect significant differences in gene composition between the control and treatment groups (**Supp. Figure 3**). We then investigated individual orthologs that could be responsible for the difference in metabolic potential. Since our preclinical data identified SCFAs and butyrate as key effectors of vancomycin effects, we first examined pathways associated with SCFAs, including glycoside hydrolases, which break down complex carbohydrates into simpler sugars in the gut^22^. KEGG pathway analysis revealed differences between control and treatment groups at each time point for glycoside hydrolase genes (**Figure 3B**). These differences are likely due to the overall decrease in members of the Bacteroidetes phylum and modulation of specific genera and species capable of performing this function. We then investigated the levels of butyrate producing genes since the most common mechanism of butyrate production in the gut is through fermenting complex carbohydrates along with three other pathways that rely on amino acid metabolism. In our cohort, the aggregated levels of genes in four butyrate producing pathways showed decreasing trends across time in the treatment cohort, and the levels of glutarate and 4-aminobutyrate (GABA) pathways were altered at the start of SBRT (**Figure 3C**). The Bcd gene, which is a bottleneck gene common to all four pathways, as well as the Buk and But genes which are terminal genes in the production of butyrate in the fermentative pathway were lower at the start of SBRT compared to baseline in the treatment group, leading to decreased butyrate producing potential of the microbiome^23^ (**Figure 3D**). Bacteria in the gut can modify primary bile acids secreted into the gut lumen into secondary bile acids through specific converting enzymes. The set of genes responsible for processing primary bile acids is defined by the bai operon, while the deconjugation of amino acids, which is a crucial step in this conversion process, is mediated by the BSH gene. In our cohort, we observed both these pathway were negatively affected by vancomycin (**Figure 3E**) ^24^.

**Figure 3:**
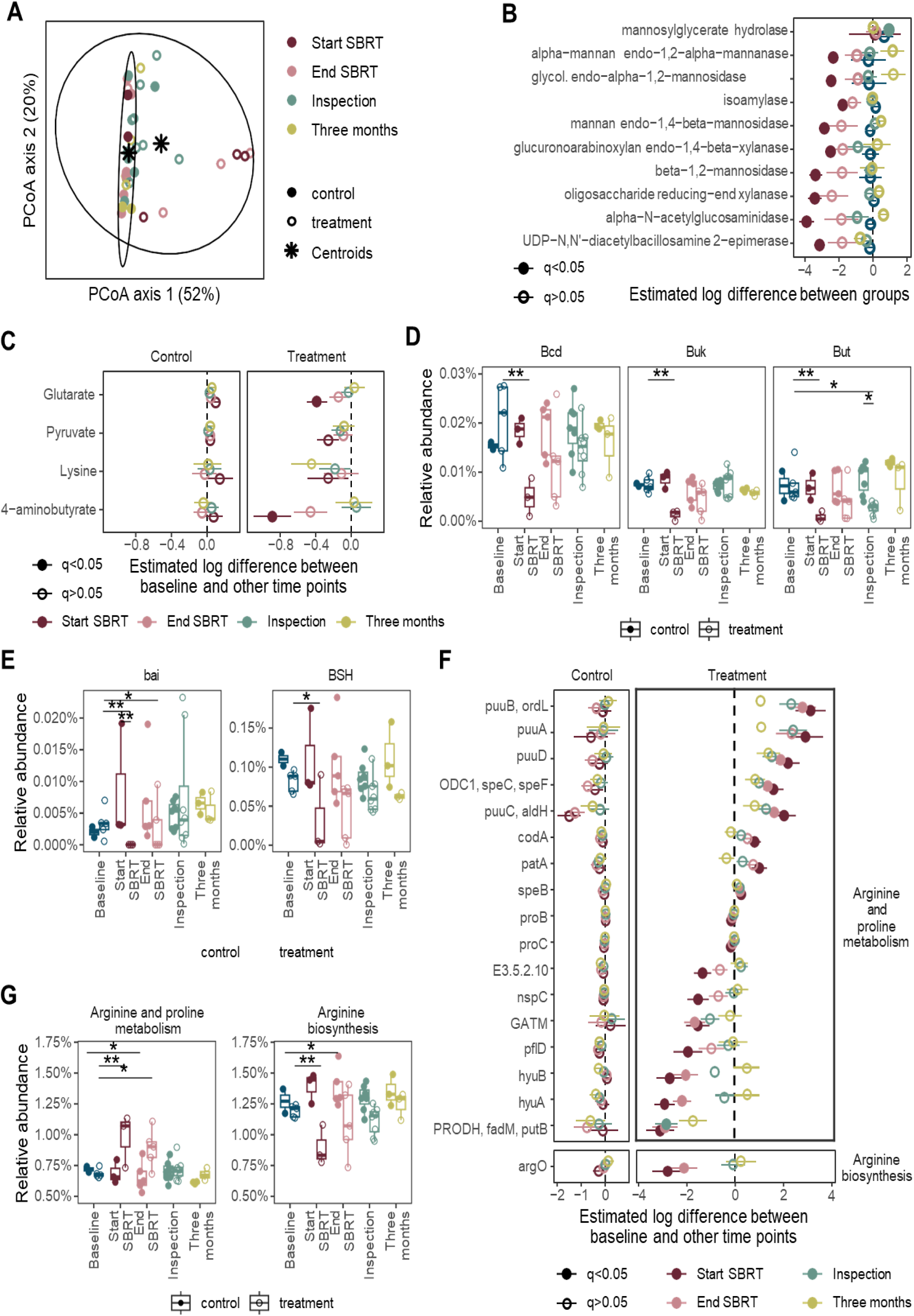
Effect of vancomycin treatment on bacterial gene abundances. A) Principal coordinate analysis on Bray-Curtis distances of gene abundances of bacteria obtained through alignment to KEGG database. B) Estimated log difference between control and treatment groups at each time point for glycoside hydrolase genes that are significantly different (linear models) C) Estimated log difference between baseline and other time points in each study group for the major butyrate production pathways (linear mixed effects models). D) Relative abundance of major genes in butyrate production by bacteria in the gut. E) Relative abundance of bile inducible operon (bai) and bile salt hydrolase (BSH) genes. G) Estimated log difference between baseline and other time points for genes that were altered in select pathways. ***<0.001, **<0.01, *<0.05.

Following up on additional metabolite analysis showing alterations the total relative abundance of the genes in arginine biosynthesis were lower and arginine metabolism were higher in the vancomycin group (**Figure 3F**). One mechanism of arginine metabolisms starts with converting arginine into ornithine^25^. The genes ornithine decarboxylase (speC/speF) responsible for converting ornithine to putrescine, genes puuA, puuB, puuC, puuD responsible for converting putrescine to 4-aminobutyrate and patA responsible for converting putrescine to 4-aminobutanal were all elevated while subjects were on vancomycin (**Figure 3G**).

### Validation of the Impact of vancomycin on gut microbiome and consequent impact on serum metabolome

Building upon our comprehensive investigation into vancomycin’s impact on the gut microbiome and its subsequent effects on metabolic KEGG pathways, we extended our research to explore the broader systemic implications of these microbiome alterations. The next phase of our study encompasses a detailed metabolomic analysis, employing Liquid Mass Spectrometry (LMS-TOF) to quantitatively and qualitatively assess the serum metabolite profiles of patients treated with vancomycin compared to control subjects. Complementing our KEGG analysis, which highlighted pathways involved in SCFA production, we observed pronounced changes in SCFAs levels following vancomycin treatment. Our results showed that vancomycin significantly diminished all tested SCFAs in stool samples: acetic acid, propionic acid, isobutyric acid, butyric acid, isovaleric acid, and hexanoic acid (**Figure 4A and supp. Figure 4A**). In plasma samples, significant reductions were detected in butyric and isovaleric acid, while acetic acid, propionic acid, isobutyric acid and hexanoic acid levels did not show significant changes (**Figure 4B**). This suggests that vancomycin’s impact on SCFAs production in the gut is mirrored to some extent in systemic circulation, particularly affecting butyric acid and isovaleric acid. The visual representation underscores the systemic effects of vancomycin on metabolite in the serum, reflecting its impact on host metabolism when used as a therapeutic agent in combination with SBRT (**Figure 4C and Supp. Table 3**). Recent research has highlighted the relevance of bile acids in cancer and immunotherapy, the efficacy of immunotherapeutic strategies^26^. In our mass spectrometry analysis, we found that specifically lithocholic acid levels was decreased (**Figure 4D**).

**Figure 4:**
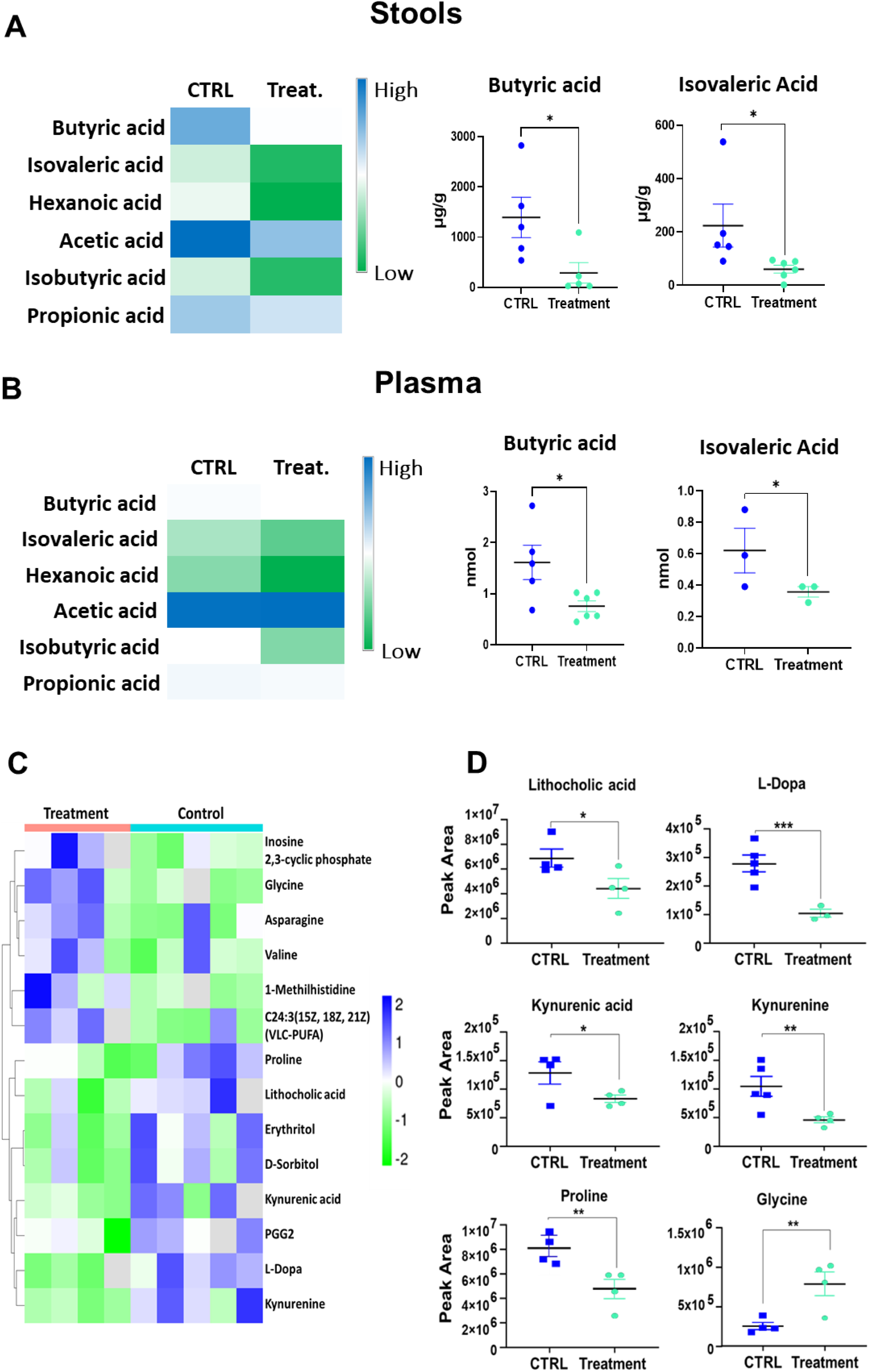
Impact of Vancomycin Treatment on Short-Chain Fatty Acids (SCFAs) and Other Metabolites in Stool and Plasma. **A.** *Stool* SCFAs: *left* heatmap, *right* display level of butyric acid and isovaleric acid in the treatment group compared to the control. **B.** *Plasma* SCFAs: *left* heatmap, *right* display level of butyric acid and isovaleric acid in the treatment group compared to the control. **C.** *Plasma* Metabolomics: Heatmap highlighting broad changes in serum/plasma metabolic profiles of patients receiving oral vacnomycin, **D.** Significant differences in specific metabolites between control and treatment groups: Lithocholic Acid, L-Dopa, Kynurenic Acid, Kynurenine, Proline and Glycine.

Our metabolomic analysis also revealed additional significant changes in various other metabolites in the serum. Notably, there was a decrease in L-Dopa levels in the treatment group compared to controls, suggesting an impact on dopamine biosynthesis pathways. Additionally, levels of kynurenic acid and kynurenine, both metabolites in the tryptophan degradation pathway, were decreased in the treatment group, indicating catabolism of tryptophan, which has immunomodulatory effects (**Figure 4D**). Furthermore, we observed a decrease of proline an increase of glycine indicating an impact on amino acids metabolism (**Figure 4D**). Other metabolite levels affected in the serum of vancomycin-treated patients were the long-chain fatty acid C24:3(15Z,18Z,21Z), inosine 2’,3’-cyclic phosphate (**Supp. Figure 4B**), suggesting broad alterations in nucleotide metabolism and fatty acid biosynthesis. Interestingly, we detected enhanced levels of 1-methylhistidine (1-MH) in the serum following vancomycin treatment. These changes could reflect alterations in gut microbial activity or dietary habits, potentially influencing metabolic processes like the absorption or clearance of 1-MH. While 1-MH itself is not known to affect the immune system directly, the broader metabolic shifts associated with its altered levels could have indirect implications for immune responses and the tumor microenvironment (**Supp. Figure 4B**).

### Linking Serum Metabolomics with PBMC Transcriptomics: Vancomycin’s Systemic and Immune Modulation

Due to clinical guidelines for early-stage NSCLCs, obtaining post-treatment biopsies was not feasible. Consequently, we expanded our research to include a detailed analysis of serum metabolomic alterations in patients treated with vancomycin. This approach allowed us to explore the broader cellular impacts of these metabolic changes by examining the effects on the transcriptome of circulating PBMCs. This investigation bridges the identified systemic metabolic changes with molecular mechanisms driving immune responses, offering a comprehensive view of vancomycin’s influence on host physiology. Through differential gene expression analysis, enhanced by Ingenuity Pathway Analysis, we observed significant transcriptomic alterations in PBMCs post-vancomycin treatment. A key finding was the substantial upregulation of the antigen presentation pathway (**Figure 5A**), echoing our preclinical findings in mice^11^ and illustrating vancomycin’s potent modulation of immune recognition processes across species, despite individual variability. To validate the sequencing results, we performed qPCR of several molecules central to antigen presentation. Confirming the RNA sequencing data, we observed a significant increase in the expression of PSMB8, TAP1, B2M, CALR, and BATF3 (Figure 5B). These molecules are crucial for the antigen presentation pathway, underscoring the impact of vancomycin on enhancing immune surveillance mechanisms^27^. To assess the potential effects on leukocyte population phenotypes, we conducted a comprehensive 16-color FACS analysis on PBMCs collected after SBRT treatment. This analysis encompassed T cells (CD45+, CD3+, CD4+, CD8+ and FoxP3), neutrophils (CD45+CD3-CD15+), dendritic cells (DCs; CD45+CD11b-CD11c+), natural killer cells (NKs; CD45+CD3-CD56+), monocytes (CD45+CD14+CD16+), and myeloid-derived suppressor cells (MDSCs; CD45+CD33+CD14/15+HLA-DR low). Despite the inherent variability among individuals, we noted a notable expansion of dendritic cells expressing HLA-DR, T cells expressing CD25 and PD1, and NK cells expressing CD25 in patients treated with vancomycin (**Figure 5C**). Cytokines play a crucial role in cancer therapy by mediating and enhancing the antitumor immune response. Specifically, Th1 cytokines are pivotal in this process^14^. Therefore, we performed several enzyme-linked immunosorbent assays (ELISAs) to detect the levels of several cytokines including IFN-β, IFN-γ, TNFα and IL-18. While the increases in IFN-β and IFN-γ levels observed in the vancomycin-treated group did not reach statistical significance, there was a discernible trend towards elevated levels, suggesting a potential enhancement of antiviral and antitumor responses (Figure 5D, top and bottom). However, most serum samples did not exhibit detectable levels of TNFα or IL-18, as shown in Supplementary Figure 5.

**Figure 5.**
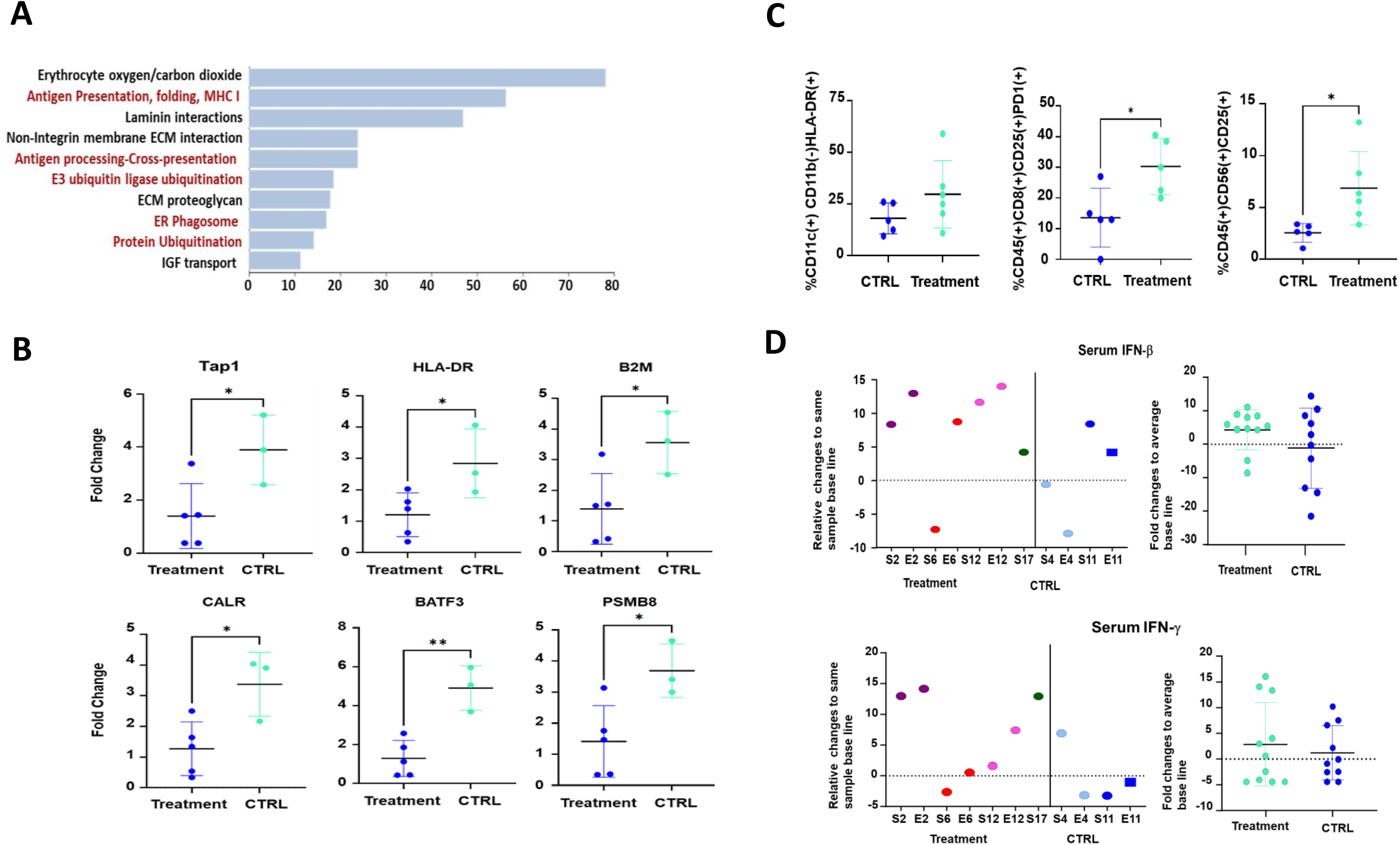
Impact of Oral Vancomycin on PBMC Transcriptome, Phenotype, and Serum Cytokine Levels. **A)** Ingenuity pathway analysis of RNA sequencing data from twelve samples collected one week after vancomycin administration, highlighting significant changes in immune-related pathways. **B)** qPCR validation shows increased expression of antigen-presenting genes in PBMCs from vancomycin-treated patients. **C)** FACS analysis reveals changes in the phenotype of immune cells within PBMCs, including DCs, T cells, and NK cells, characterized by increased expression of activation markers such as HLA-DR, PD-1, and CD25. **D)** Serum levels of Th1 cytokines IFN-β and IFN-γ, showing treatment-induced changes compared to baseline.

In conclusion, our integrative approach, combining serum metabolomics and PBMC transcriptomics, underscores vancomycin’s systemic effects and its specific influence on immune modulation. The drug not only alters serum metabolite profiles but also impacts the immune system at the cellular and molecular levels, particularly enhancing antigen presentation pathways and cytokine responses, thereby potentially boosting antitumor immunity.

## Discussion

The findings from this study highlight the impact of vancomycin-induced alterations in the gut microbiome on host metabolism and immune response, emphasizing their relevance to enhancing the efficacy of RT for patients with early-stage NSCLC. Vancomycin treatment reshapes the gut microbiota, leading to a substantial reduction in microbial diversity—a pivotal change that sets the stage for broader systemic effects. This microbial imbalance, characterized predominantly by a decrease in Bacteroidetes and an increase in Proteobacteria, signals a significant disruption that extends beyond the gut to influence systemic metabolic processes^15,28^.

Building on these microbial changes, our study provides insights into the metabolic responses following vancomycin treatment in the context of SBRT. Using the KEGG database, we traced the treatment’s impact on key metabolic pathways, revealing a decrease in the activity of glycoside hydrolases. Mirroring the preclinical studies this enzymatic reduction underpins the disruption of SCFAs production—metabolites known for their anti-inflammatory properties^11,16^. The decline in SCFAs producers, particularly from the Lachnospiraceae and Ruminococcaceae families^17,29^, not only alters the metabolic landscape but also suggests a shift towards a pro-inflammatory state, potentially recalibrating the immune system’s response to tumor RT treatment. The aggregated levels of genes involved in butyrate-producing pathways showed decreasing trends over time in the treatment group, with significant alterations in the GABA pathways during vancomycin treatment^30^. GABA receptors are expressed on various immune cells, including T cells, where their activation can influence T cell proliferation and cytokine production, potentially modulating the anti-tumor immune response by reducing pro-inflammatory cytokines like IFN-γ and TNF-α^31–33^. In cancer therapy, GABA’s anti-inflammatory properties could counteract the pro-inflammatory environment required for ICI and CAR-T therapy impacting their efficacy^34^. Vancomycin treatment also resulted in reduced levels of genes associated with converting primary to secondary bai operon and BSH gene, suggesting a diminished capacity for bile acid modification. Additionally, arginine and purine metabolism were influenced by lower relative abundance of genes responsible for arginine biosynthesis and elevated levels of genes involved in arginine metabolism.

The immune modulation observed in this study is intricately tied to the microbial and metabolic alterations. Enhanced activation of immune cells, as evidenced by flow cytometry findings, shows an uptick in the APCs functionality of dendritic cells, as well as T cells and NK cells expressing markers such as CD25 and PD1^35–37^. These changes suggest an environment primed for a heightened antitumor response, potentially facilitated by altered metabolic cues stemming from vancomycin treatment. This link is further supported by shifts in cytokine profiles, where trends in increased IFN-β and IFN-γ levels underscore the antitumor response^14^.

In line with our and others previous works the metabolomic analysis using MS-TOF revealed significant reductions in key SCFAs in stool samples, with notable reductions of only butyric acid and isovaleric acid in serum, while other SCFAs like acetic, propionic, and isobutyric acids remained unaffected. SCFAs, particularly butyrate, propionate, and valerate, exhibit potent anti-inflammatory effects by modulating the function of various immune cells^38,39^. These effects include enhancing Treg function^29^, suppressing pro-inflammatory cytokine production in T cells^40^, inhibiting DCs APCs and IL-12 secretion^41,42^, and macrophages^43^, and improving gut barrier integrity^44^. In the context of RT for cancer treatment, an inflammatory response that enhances APC functions and T-cell activity is crucial for effective tumor eradication. Our study observed a reduction in SCFAs levels, which may be beneficial in this therapeutic setting. Lower SCFA levels could potentially increase immune system alertness to threats and strengthen antitumor immune responses. This reduction might promote a pro-inflammatory environment, activating the immune system more effectively and potentially enhancing the overall efficacy of RT. Although this approach might seem to contrast with the beneficial role of SCFAs in managing autoimmune diseases—where reducing inflammation is crucial—it makes sense in the context of cancer therapy, highlighting the context-dependent effects of these metabolites. While SCFAs have traditionally been recognized for their anti-inflammatory properties^45,46^, recent research suggests a more nuanced role in immune modulation, with specific SCFAs influencing different leukocyte populations. For instance, butyrate specifically suppresses DC functions^41^ while enhancing T cell responses to infection and cancer^47^. Similarly, valerate improves T cell function^19^ while inhibiting the activation of neutrophils and monocytes^48^. These findings underscore the complex interplay between the gut microbiome, SCFAs production, and the immune system, highlighting the potential of antibiotic treatment to modulate these metabolites and their impact on host immunity and metabolism. In line with previous work and our pathway analysis, vancomycin administration affected bile acid metabolism by decreasing levels of lithocholic acid ^16^. Although our metagenomic sequencing data did not specifically target the bacteria involved, a possible explanation for these effects is that vancomycin reduces the population of bacteria that convert chenodeoxycholic acid into lithocholic acid, particularly Clostridia and other bacterial genera involved in 7α-dehydroxylation, leading to decreased LCA production^49^. Lower lithocholic acid levels result in reduced activation of vitamin D receptor (VDR) and pregnane X receptor (PXR), decreasing immune suppression and anti-inflammatory effects^50^. The decrease in LCA may contributes to a less immune-suppressive environment, potentially leading to heightened immune activation.

The observed toxicity profile suggests that vancomycin treatment is well-tolerated, with only mild to moderate side effects such as diarrhea, nausea, and fatigue reported, and no severe adverse events. This encouraging safety profile enhances the potential for integrating vancomycin into existing cancer treatment regimens, offering a promising avenue for enhancing therapeutic efficacy with manageable toxicity. While the effects of vancomycin on the gut microbiome and immune modulation are evident, it is prudent to consider certain variables that might have influenced these outcomes, such as variations in patient diet, concurrent medications, and underlying health conditions. Recognizing these factors allows for a more nuanced interpretation of the results and helps delineate areas for further investigation. Although the sample size of our study was modest, the consistent patterns observed affirm the validity of the underlying biological mechanisms previously identified in preclinical models^11,51^. This alignment underscores the translational potential of our findings and supports the hypothesis that microbial modulation can significantly impact immune responses and cancer outcomes. To build on these promising results and overcome the limitations posed by the sample size, future studies with larger cohorts are essential.

Despite these challenges, our study makes a significant contribution to the field by demonstrating the feasibility of modulating the gut microbiome to enhance immune-mediated cancer therapies. The insights gained here lay the groundwork for future interventions that could potentially transform cancer treatment paradigms.

In essence, vancomycin and RT work through distinct but complementary pathways to significantly enhance the immune system’s ability to combat cancer effectively. Vancomycin modulates the systemic environment via adjustments in the gut microbiome, while RT directly affects tumor cells and the local tumor environment^28,52^. This dual approach not only leverages enhanced immune activation and increased antigen availability but also capitalizes on the synergy between systemic immune readiness and localized immune stimulation, creating a scenario where the immune system is better equipped to recognize and attack cancer cells. The observed improvements in OS and PFS among patients treated with vancomycin suggest that integrating microbiome-targeting strategies with conventional therapies like RT can substantially improve clinical outcomes. Vancomycin’s established safety profile and broad availability make it an appealing therapeutic option, particularly in scenarios where more complex treatments may be impractical due to higher toxicities or resource constraints.

This strategy underscores the innovative potential of using established, widely accessible treatments like vancomycin alongside RT. It offers a cost-effective, low-toxicity method to enhance cancer therapy. Future research should aim to refine our understanding of how to best harness the therapeutic benefits of targeting the gut microbiome to further enhance cancer treatment outcomes, ensuring that microbiome-modulating therapies are used effectively and safely.

### Statement of Contribution

S.F. led the clinical trial and assisted in drafting the manuscript. C.F. performed most of the key laboratory experiments and helped in drafting the manuscript. C.T. and K.B. preformed shotgun sequence and analyzed the metagenomic data. R.O’C., B.V., A.A.S., T.Q. were instrumental in performing and interpreting the mass spectroscopy data. D.A. carried out the RNA Ingenuity Pathway Analysis and helped in drafting the manuscript. N.K. performed sample collection, DNA extraction and logistic for samples sequencing. G.S. and A.H. conducted RNA sequencing deconvolution, performed differential gene expression analysis. S.S analyzed the mass spectroscopy data and prepared the heatmaps. Team members C.F., L.C., N.Y.-R., K.C., B.L., J.B., supported the development of the clinical trial and participated in the revision of the paper. A.B. initiated the clinical trial and contributed to the drafting of the clinical protocol.

A.M. and S.L. provided significant support in manuscript revision and data interpretation. C.K. was involved in the conceptualization of the idea and financial support. E.B.-J. was responsible for writing the clinical protocol and assisting in manuscript drafting. A. F. conceptualized the study, help the protocol writing, led the laboratory experiment and analyses, wrote the manuscript.

## Data Availability

All data produced in the present work are contained in the manuscript https://www.ncbi.nlm.nih.gov/sra https://www.ncbi.nlm.nih.gov/geo/

## Acknowledgments

We would like to thank the NIH/NCI for financial support through grant 1R01CA219871-01A1. We also extend our gratitude to Dr. M. Uribe-Herranz and Dr. S. Beghi for their assistance with sample collection and aliquoting. We especially acknowledge Norma Carretta for her invaluable career guidance and support, and we remember her with great respect following her recent passing. Most importantly, the study team thanks all the patients who contributed their time, samples, and data to this research.

