## Supplemental Materials and Methods for "“Immune System Modulation with Oral Vancomycin in combination with Stereotactic Body Radiotherapy (SBRT) for medically inoperable Early-Stage Non-small Cell Lung Cancer”"

*Metagenomic sequencing*

DNA was extracted from 200 mg of stool and quantified using the Quant-iTTM PicoGreen dsDNA assay kit (Thermo Fisher Scientific). Shotgun libraries were generated from 7.5 ng DNA using Illumina DNA Prep Library Prep kit and IDT for Illumina unique dual indexes at 1:4 scale reaction volume. Library success was assessed by Quant-iT PicoGreen dsDNA. An equal volume of library from each sample was pooled and sequenced using a 300 cycle Nano kit on the Illumina MiSeq. Libraries were then repooled based on the demultiplexing statistics of the MiSeq Nano run. Final libraries were run on the Agilent BioAnalyzer to check the size distribution and absence of additional adaptor fragments. Libraries were sequenced on an Illumina Novaseq 6000 v1.5 flow cell, producing 2x150 bp paired-end reads. Extraction blanks and nucleic acid-free water were processed along with experimental samples to empirically assess environmental and reagent contamination. A laboratory-generated mock community consisting of DNA from Vibrio campbellii and Lambda phage were included as a positive sequencing control.

*Bioinformatics processing*

Resulting fastq files from metagenomic sequencing were analyzed using Sunbeam18. Quality control steps were performed by removing adapters, trimming low quality reads and removing low complexity and host-derived sequences per default workflows in Sunbeam. The abundance of bacteria was estimated using Kraken19. Reads were mapped to the KEGG20,21 database to estimate the abundance of bacterial gene orthologs and to curated databases of genes involved in butyrate production22. Sample similarity was be assessed by Bray-Curtis distance.

*Metabolomic analysis MS-TOF*

For metabolomic profiling, we performed liquid chromatography-mass spectrometry (LC/MS) on serum samples from 9 patients, divided as 5 from the SBRT group and 4 from the SBRT plus vancomycin group. Metabolites were extracted by adding 300 µL of 0°C cold methanol to 100 µL of serum, vortexing, resting on ice for 10 minutes, and then centrifuging at 21,000 G for 15 minutes at 0°C. The supernatant was subjected to solid phase extraction using an Agilent Positive Pressure Monifold 48 Processor equipped with Agilent Captiva EMR-Lipid cartridges. The cleaned supernatants were again centrifuged, and 100 µL aliquots were transferred to autosampler vials equipped with 250 µL polyethylene inserts for LC analysis.

Analysis was conducted using a 6546 QTOF LC/MS system (Agilent Technologies, CA, USA) employing hydrophilic interaction chromatography (HILIC) on an InfinityLab Poroshell 120, HILIC-Z, 2.1 × 150 mm, 2.7 micron LC column (Agilent Technologies, catalog no. 6546). Metabolites were eluted with a nonlinear gradient of LC-MS grade acetonitrile (Supelco LiChrosolv, mobile phase B) and 10 mM ammonia acetate in HPLC water with 5 µM medronic acid and ammonia hydroxide (phase A, pH=9.30) at a flow rate of 400 µL/min. MS data acquisition was performed in negative mode from 50 to 1100 Da at one scan/s in both centroid and profile modes in 4-GHz high-resolution mode, calibrating using negative-ion reference masses of 112.985587 and 1033.988109 m/z (Agilent Technologies).

Instrument performance was monitored by analyzing quality control samples consisting of a metabolite extract of pooled serum samples at the start of each day and after every six samples. U-13C labeled metabolite standard mixes and yeast extract compounds (Cambridge Isotope Laboratories, MA, USA) were utilized as internal standards. Data were analyzed using MassHunter Profinder version 10.0 software (Agilent Technologies, CA, USA).

*Heatmap procedure.*

A total of nine samples were analysed, divided into two groups: four samples in triplicate treated with vancomycin and five untreated control samples in triplicate. The relative levels of specific metabolites were measured for each sample using mass spectrometry, see above. The dataset included both individual metabolite values for each sample and group averages ("vancomycin average" and "control average"). Raw data were pre-processed to correct column naming inconsistencies and ensure uniform formatting for analysis. Heatmaps were generated using the R programming language (version 4.4.0) and the pheatmap package to visualize both the individual variations across samples and the overall differences between treated and control groups. Hierarchical clustering was applied to identify patterns and relationships between metabolites and samples. The top 10 and top 30 metabolites were selected based on their statistical significance, as determined by a two-tailed t-test comparing the treated and control groups. Metabolites with the smallest p-values were prioritized for inclusion in the heatmaps.

*Quantitative RT-PCR*

The relative quantification of the expression levels of selected genes was carried out by qPCR. Total RNA from cells and tissues was extracted using TRIzol reagent (Invitrogen, 15596018) according to the manufacturer's instructions. Reverse transcription and RT-PCR reactions were carried out using the High Capacity cDNA Reverse Transcription Kit (Thermo Fisher Scientific, 4368814) and TaqMan Gene Expression Master Mix (Thermo Fisher Scientific, 4369016) according to the manufacturer's instructions. Runs were performed using the QuantStudio 6 Flex Real-Time PCR System (Thermo Fisher Scientific). All TaqMan primers were purchased from Thermo Fisher Scientific. Housekeeping gene Rn18s (Hs99999901_s1), IL12A (Hs01073447_m1), TAP1 (Hs00388675_m1), HLA-DRA (Hs00219575_m1), PSMB9 (Hs00160610_m1), B2M (Hs00187842_m1), CALR (Hs00189032_m1), PSMB8 (Hs00544758_m1), HLA-A (Hs01058806_g1), BATF3 (Hs00232744_m1), TBX21 (Hs00894392).

*Flow cytometry*

Dead cells from patients PBMCs were removed with the Dead Cell Removal Kit (Miltenyi Biotec 130-090-101). Alive cells were subjected to flow cytometry on a FACS Fortessa flow cytometer using BD FACS Diva software (BD Biosciences) and data were analyzed using FlowJo version 10.8.1. LIVE/DEAD Fixable Aqua Dead Cell Stain (Invitrogen, L34966) was used to gate living cells. The following mAbs against human markers were used to phenotype the immune cells: αHLA-DR (clone L243, Biolegend), αCD25 (m-a251 (ruo), BD Horizon) αCD8a (clone RPA-T8, Biolegend), αCD15 (clone SSEA-1, Biolegend), αPD1 (clone NAT105, Biolegend), αCD14 (clone M5E2, Biolegend), αCD68 (clone y1/82a, Biolegend), αCD3 (clone HIT3a, Biolegend), αPDL-1 (clone 29E2A3, Biolegend), αCD11c (clone Bu15, Biolegend), αCD45 (clone 2D1, Biolegend), αCD56 (clone NCAM, Biolegend), αFoxP3 (clone 259D/C7 (RUO), BD Pharmingen), αCD4 (clone RPA-T4, Biolegend), αCD33 (clone P67.6, Biolegend), αCD123 (clone 9F5 9 (ruo), BD OptiBuild).

*ELISA*

The levels of IFNγ, IFNβ, TNFα, IL-18, from serum/plasma of patients were evaluated using the Human DuoSet ELISA Kit (R&D Systems, DY285B for IFNγ, DY814-05 for IFNβ, DY210-05 for TNFα, DY318-05 for IL-18) and following the manufacturer's instructions.

*Total RNA seq.*

Total RNA from 5 million of PBMCs was extracted using Trizol and sent to Genewiz (N.J. USA) for total RNA sequencing.

*RNA sequencing data analysis*

Raw RNA-Seq datasets were quality checked and pre-processed using FastQC (www.bioinformatics.babraham.ac.uk/projects/fastqc/) and Cutadapt. Alignment of the trimmed sequencing reads was conducted against the human genome (version GRCh38) using the STAR aligner. Gene-level quantification was performed using RSEM and expected counts for each gene were extracted for downstream analysis. Median-by-ratio normalization was applied to RSEM results and differential expression analysis was conducted using EBSeq, setting the number of the Expectation Maximization algorithm iterations to 5. Differentially expressed (DE) genes were filtered based on the False Discovery Rate (FDR <= 0.05).

*Pathway Analysis*

We used Ingenuity Pathway Analysis to interpret the genes of interest that are involved in the biological process and pathways. An enrichment chart was generated using the pathfindR package in statistical programming language R version 4.3.0. Transcriptomic results were normalized on a natural logarithmic scale and a false discovery rate of 10% of set for pathway analysis. Multiple hypothesis testing correction for FDR-adjusted p-values was performed using the Benjamini-Hochberg method23.
