## Supplementary material for "“Immune System Modulation with Oral Vancomycin in combination with Stereotactic Body Radiotherapy (SBRT) for medically inoperable Early-Stage Non-small Cell Lung Cancer”": Supplemantal Figures and Tables

### Supplemental Figure 1

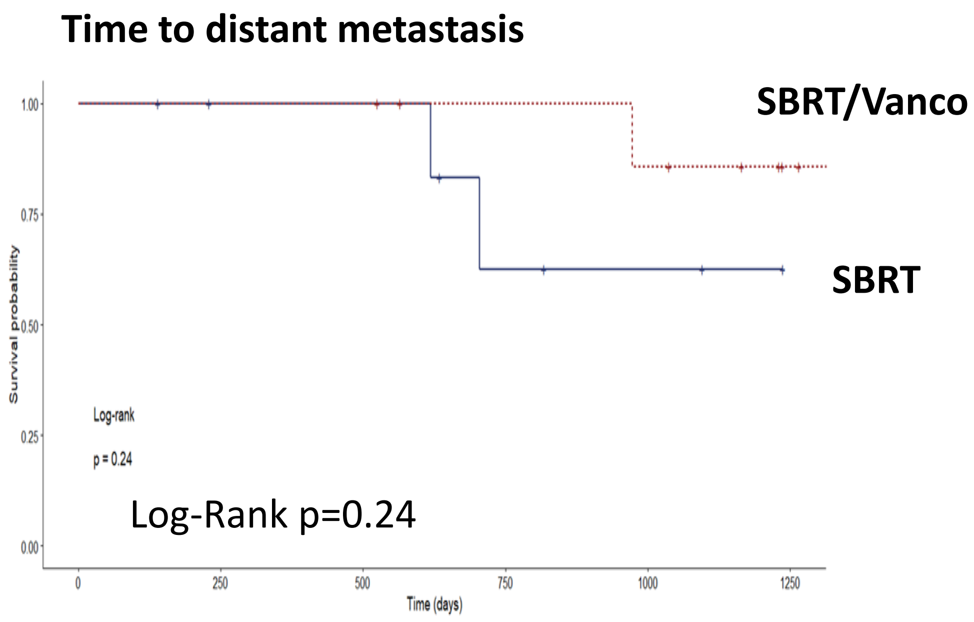


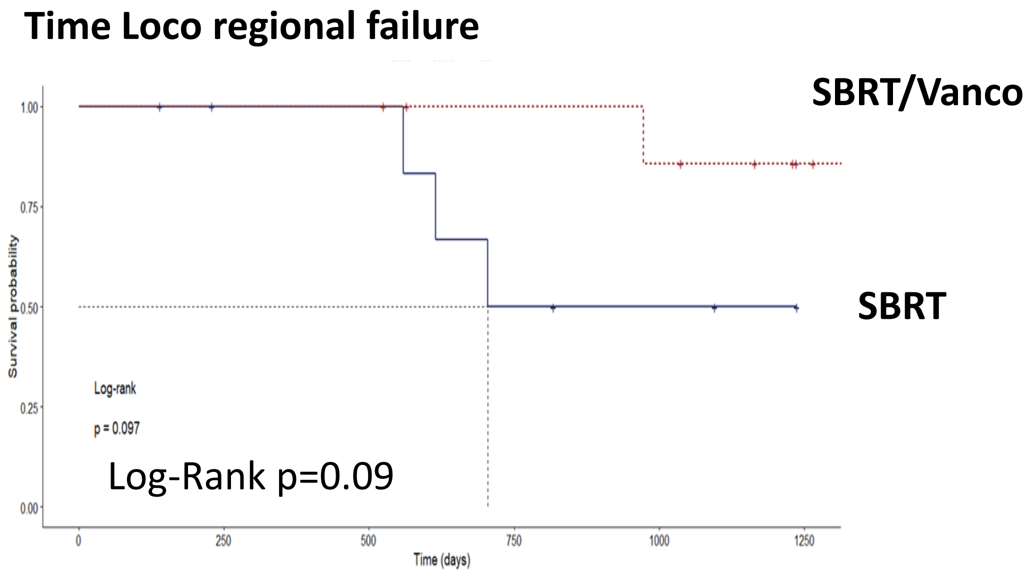


**Supplemental Figure 1**
**Top.** time to distant metastasis **bottom.** local and locoregional failures

### Supplemental Figure 2

A

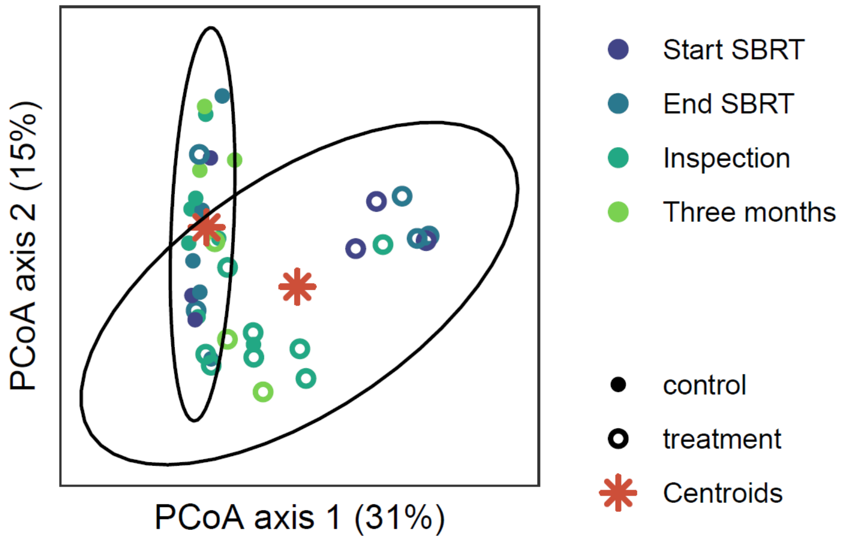


B

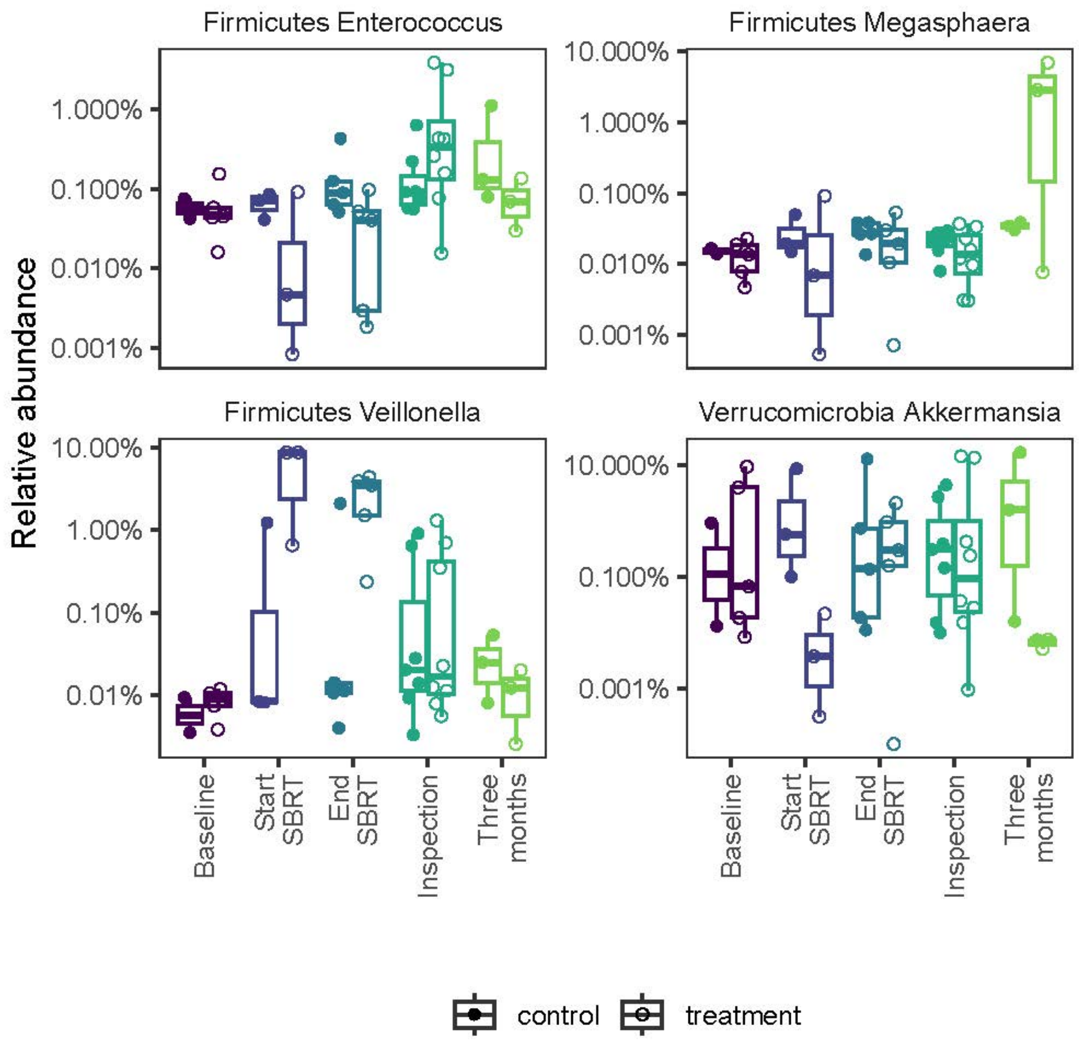


**Supplemental Figure 2. A**) Cumulative principal coordinate analysis on Bray-Curtis distances of taxonomic abundances of bacteria. **B**) Relative abundance of the genra Akkermansia, Veillonella, Megasphaera and Enterococcus. Differences across time were tested using linear mixed effects models and differences between control and treatment groups were tested using linear models. ***<0.001, **<0.01, *<0.05. D) Mean of center log ratio (clr) transformed abundances at each time point. The stars mark the bacteria that are different between the control and treatment groups for that time point using linear models.

### Supplemental Figure 3

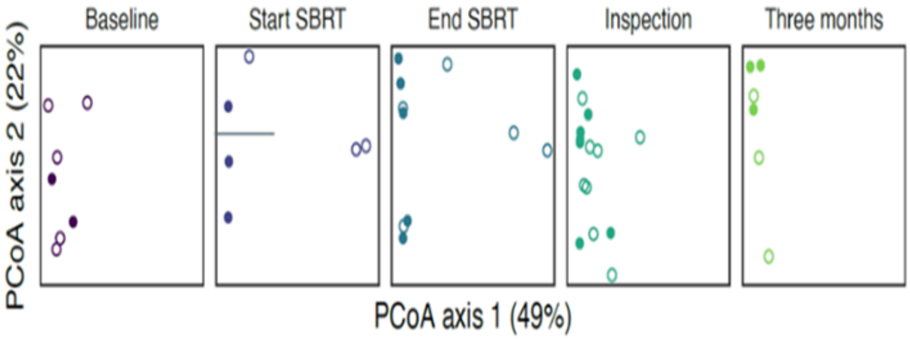


**Supplemental Figure 3.** Principal coordinate analysis (PCoA) on Bray-Curtis distances, based on taxonomic abundances of bacteria sampled at various times from treated patients and control.

#### Supplemental Figure 4

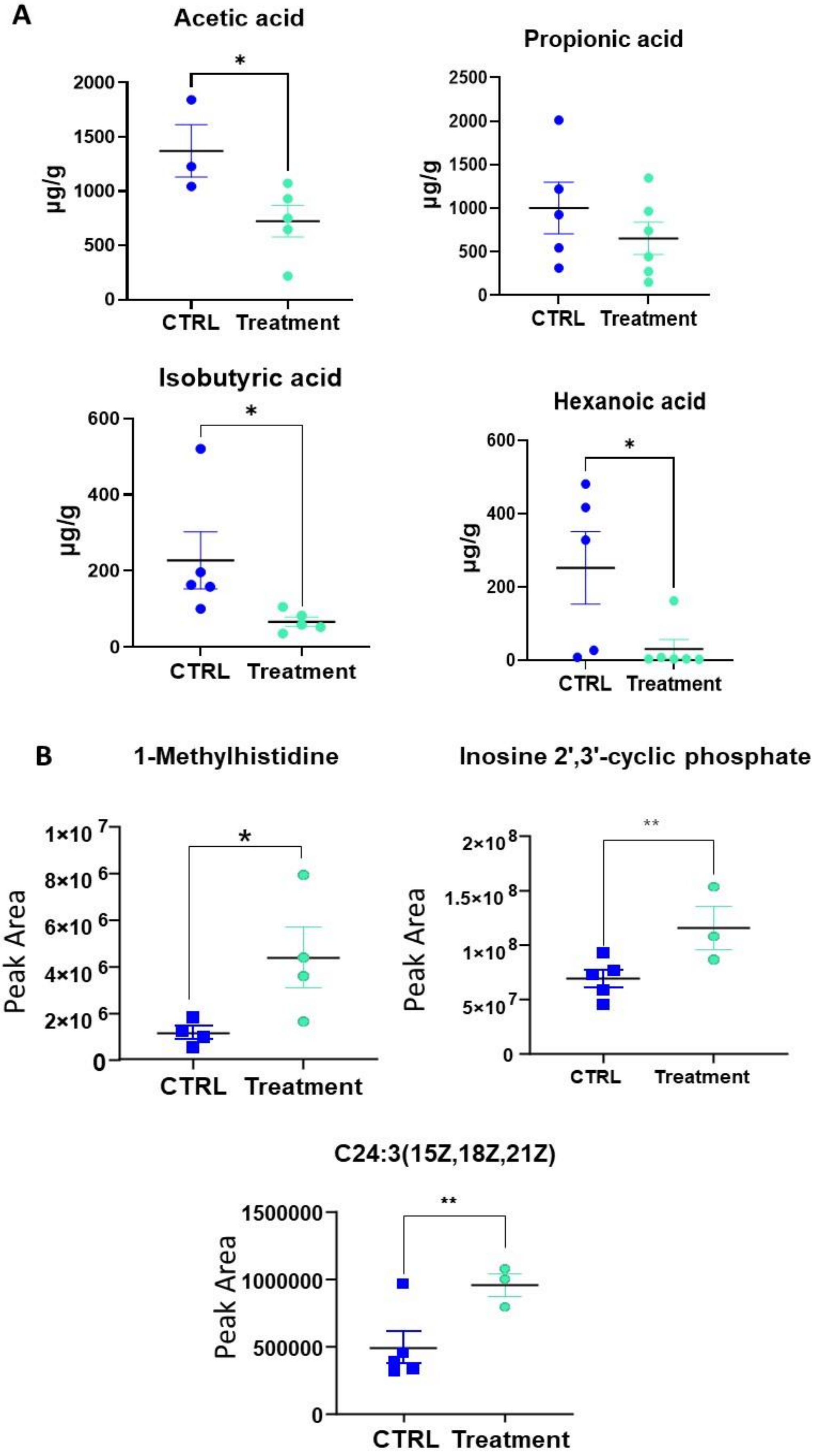


**Supplemental Figure 4: A)** Additional short-chain fatty acids (SCFAs) in stool samples from patients treated with vancomycin versus controls. **B**) Additional metabolites in serum samples.

### Supplemental Table 3

| | | | | SBRT+VANCO | | | | | | SBRT | | | | | | | |
|---|---|---|---|---|---|---|---|---|---|---|---|---|---|---|---|---|---|
| Metabolite/Isobar | Mass (a | Found | | S2 avarage | S3 average | S12 avarage | S16 avarage | avarage of S2,3,12 | | S4 avagare | S5 avarage | S10 avarage | S11 avarage | S15 avarage | avarage S4,5,10,11,1 | | P value |
| Succinic acid | 118 | 13 | | 7992490.7 | 11164744 | 5389904 | 4466050 | 7253297 | | | 22412147 | 8848723 | 6743250 | | | | 0.28 |
| Succinic semialdehyde | 102 | 17 | | 28441760 | 26444440 | 33949639 | | 3E+07 | | 15698693 | 80864157 | 19696418 | 38228316 | 12383460 | 33374209 | | 0.832 |
| D-Alanine | 89.05 | 14 | | 1466037 | 989559 | 637786 | 544825 | 909552 | | 536927 | 1071116 | | 899972 | | 836005 | | 0.803 |
| D-Glucose 6-phosphate | 260 | 13 | | 648581 | 286915 | 308597 | 87899 | 332998 | | 437080.3 | 146713 | | 201377 | | 261723.44 | | 0.668 |
| Malic acid | 134 | 18 | | 6851361 | 1439291 | 1167567 | 566528 | 2506187 | | 1285702 | 8932907.3 | 4655058 | 2927343 | 1125427 | 3785287.5 | | 0.557 |
| Lactic acid | 90.03 | 16 | | 134137451 | 149568627 | 134000070 | 43847468 | 1.2E+08 | | 61028687 | 395217386 | 86001429 | 89081725 | 98671285 | 146000102 | | 0.692 |
| Alanine | 89.05 | 14 | | 1466037 | 989559 | 637786 | 544825 | 909552 | | 536927 | 1071116 | | 899972 | | 836005 | | 0.803 |
| Serine | 105 | 18 | | 1536044.8 | 1343555.3 | 1108519 | 1353170 | 1335322 | | 1289269 | 1097044.7 | 931909 | 954320 | 1360861 | 1126680.7 | | 0.138 |
| Citric acid | 192 | 14 | | 5502523.7 | 10165528 | | 2918333 | 6195462 | | 1399612 | 9811218.3 | | 14189306 | 10650915 | 9012762.8 | | 0.477 |
| Valine | 117.1 | 18 | | 10400593 | 13678890 | 11287937 | 7733224 | 1.1E+07 | | 6363255 | 8607741.3 | 13276841 | 8936172 | 8743777 | 9185557.2 | | 0.373 |
| Ornithine | 132.1 | 18 | | 1754845 | 1261277 | 1281415 | 731276 | 1257203 | | 1025813 | | 2256899 | 1584768 | 1118013 | 1496373 | | 0.521 |
| Glycine | 75.03 | 18 | | 1871592.8 | 1169088 | 1235187 | 713853 | 1247430 | | 1109055 | 656006.33 | 1221393 | 1244843 | 1137713 | 1073802.1 | | 0.496 |
| Isoleucine | 131.1 | 18 | | 6822037.5 | 8375982 | 6731574 | 5045678 | 6743818 | | 3701806 | 7008894.7 | 10058255 | 5975272 | 6153388 | 6579523.2 | | 0.904 |
| Cysteine | 121 | 16 | | 1437004.8 | 205496 | 205840 | 105731 | 488518 | | 329176.7 | 424425 | 867723 | 333008 | | 488583.17 | | 1 |
| Proline | 115.1 | 18 | | 5886043 | 5886043 | 4576224 | 2604002 | 4738078 | | 2944843 | 6820147 | 8599685 | 9403279 | 7172424 | 6988075.6 | | 0.016 |
| Threonine | 119.1 | 18 | | 14772972 | 7683246.7 | 7747189 | 4704532 | 8726985 | | 7836165 | 4394437.3 | 8050480 | 7923919 | 6891762 | 7019352.7 | | 0.429 |
| Tyrosine | 181.1 | 18 | | 3722162.5 | 4401041.7 | 2881022 | 1437182 | 3110352 | | 1205455 | 2188192.7 | 4149825 | 3323395 | 2985176 | 2770408.7 | | 0.683 |
| Tryptophan | 204.1 | 13 | | 3344295 | 4264480.7 | | 1393214 | 3000663 | | 1345135 | 2007955 | 4244427 | 2669596 | 2785189 | 2610460.4 | | 0.677 |
| 1-Methylhistidine | 169.1 | 16 | | 7929006 | 4378317 | 1642183 | 3603095 | 4388150 | | 1243897 | 1840849 | | 519063 | 983254 | 1146766 | | 0.052 |
| Asparagine | 132.1 | 18 | | 1308547 | 1488937.7 | 1626477 | 1004166 | 1357032 | | 889598.3 | 872447 | 1671246 | 847792 | 1220590 | 1100334.7 | | 0.271 |
| Asparagine | 132.1 | 18 | | 1308547 | 1488937.7 | 1626477 | 1004166 | 1357032 | | 889598.3 | 872447 | 1671246 | 847792 | 1220590 | 1100334.7 | | 0.271 |
| Ornithine | 132.1 | 18 | | 1754844.5 | 1261276.7 | 1281415 | 731276 | 1257203 | | 1025813 | 600521 | 2256899 | 1584768 | 1118013 | 1317202.8 | | 0.875 |
| Citrulline | 175.1 | 18 | | 5561175.5 | 3675821.3 | 3853181 | 1935743 | 3756480 | | 3105828 | 1722488 | 6662252 | 4473034 | 2922874 | 3777295.1 | | 0.986 |
| Phenylethylamine | 121.1 | 16 | | 1905364 | 2141936 | 4166399 | 3552732 | 2941608 | | 2464369 | 1787463 | 4092190 | | 4128482 | 3118126.1 | | 0.833 |
| Creatine | 131.1 | 16 | | 242510.33 | 449864.67 | 430528 | 270412 | 348329 | | 126998.3 | 418813 | 516939 | | 572881 | 408907.83 | | 0.61 |
| Butyric acid | 88.05 | 18 | | 6957746.3 | 2111782.7 | 2322553 | 650285 | 3010592 | | 291760 | 6628407.7 | 1487281 | 5315202 | 1368872 | 3018304.5 | | 0.997 |
| Isobutyric acid | 88.05 | 18 | | 9613228.3 | 8969292.3 | 8600375 | 7642626 | 8706380 | | 5347480 | 16628846 | 12535537 | 9552687 | 7351347 | 10283179 | | 0.511 |
| Glycerol | 92.05 | 18 | | 627303 | 290218 | 428260 | 152037 | 374455 | | 115447.7 | 537365.33 | 433388 | 400292 | 313577 | 360014 | | 0.908 |
| Serotonin | 176.1 | 15 | | 547801.25 | 943744.33 | 430528 | | 640691 | | 422095 | 461526.5 | 550822 | 974856 | 880449 | 657949.7 | | 0.701 |
| Isocitrate | 192 | 14 | | 5502523.7 | 10165528 | 2322553 | 2918333 | 5227234 | | 1399612 | 9811218.3 | | 14189306 | 10650915 | 9012762.8 | | 0.477 |
| Oxaloacetate | 132 | 16 | | 5485595.5 | 9550090.5 | 8600375 | 3192858 | 6707230 | | 1563232 | 9817843.3 | | 11494666 | 10194692 | 8267608.2 | | 0.511 |
| Indole | 117.1 | 16 | | 8611882.3 | 834225.5 | 2038898 | | 3828335 | | 4555330 | 23371409 | 14012896 | 6952829 | 10644052 | 11907303 | | 0.138 |
| L-Dopa | 197.1 | 15 | | 95677.75 | 131409 | 84633 | | 103907 | | 194632 | 365043 | 246075 | 304998 | 277854 | 277720.4 | | 0.005 |
| Taurodeoxycholate | 499.3 | 15 | | 208890.5 | 842510 | 340668 | 530332 | 480600 | | 264340 | 271610.5 | 323421 | 332946 | 427451 | 323953.7 | | 0.126 |
| 3-Methyl-L-histidine | 169.1 | 16 | | 7929006 | 4378317.3 | 1642183 | 3603095 | 4388150 | | 1243897 | 1840848.7 | | 519063 | 983254 | 1146765.6 | | 0.052 |
| Ketovaleric acid | 116 | 17 | | 1451335 | 671636 | 499573 | 519727 | 785568 | | 798313.3 | 1064444 | 521338 | 430844 | 592057 | 681399.27 | | 0.672 |
| N-Methyl-a-aminoisobutyric | 117.1 | 18 | | 10400593 | 13678890 | 11287937 | 7733224 | 1.1E+07 | | 6363255 | 8607741.3 | 13276841 | 8936172 | 8743777 | 9185557.2 | | 0.373 |
| Kynurenine | 208.1 | 10 | | 494000 | 569452 | 321309 | 453900 | 459665 | | 907575.3 | 1353889.7 | 895116 | 548423 | 1506309.3 | 1042262.7 | | 0.023 |
| Kynurenic acid | 189 | 13 | | 90027 | 97093.333 | 74756 | 70170 | 83012 | | 151012 | 141435 | 70612 | 150645 | 150645 | 128426 | | 0.019 |
| PGA2 | 334.2 | 15 | | 2963561 | 183313 | 2067577 | 967871 | 1545581 | | 1344732 | 1406472 | 187195 | | | 979466.33 | | 0.432 |
| PGA3 | 332.2 | 16 | | 1106003.5 | 427513.5 | 443039 | | 658852 | | 419367 | 845651.5 | 748968 | 611984 | 698811 | 664956.3 | | 0.972 |
| PGD2 | 352.2 | 13 | | 2864474 | 899614.33 | 1588366 | 731579 | 1521008 | | 1407606 | | 1036029 | | 1010688 | | | 0.554 |
| PGG2 | 368.2 | 16 | | 215962.67 | 230709.33 | 200124 | 63008 | 177451 | | 279723.7 | 267739 | 221252 | | 294911 | 265906.42 | | 0.047 |
| D-Lactic acid | 90.03 | 16 | | 134137451 | 149568627 | 134000070 | 43847468 | 1.2E+08 | | 61028687 | 395217386 | 86001429 | 89081725 | 98671285 | 146000102 | | 0.658 |
| Adipic acid | 146.1 | 13 | | 409810.5 | 375727 | 548195 | | 444578 | | | 323557.33 | 221816 | 932805 | | | | 0.843 |
| D-Sorbitol | 182.1 | 18 | | 2207973.5 | 4062745.3 | 1175406 | 1743641 | 2297441 | | 5678067 | 3131858 | 4363757 | 2211389 | 5185585 | 4114131.1 | | 0.058 |
| Sucrose | 342.1 | 15 | | 800460 | 572876.67 | 758149 | 340229 | 617929 | | 460945.7 | 3031567.5 | 499026 | 877083 | 488307 | 1071385.8 | | 0.402 |
| Mevalonic acid (MVA) | 148.1 | 18 | | 28689831 | 12912827 | 10344267 | 6303806 | 1.5E+07 | | 9635337 | 29914592 | 22632439 | 17809129 | 8433847 | 17685069 | | 0.596 |
| Chondroitin | 379.1 | 12 | | 246090.5 | 75015.667 | 59909 | | 127005 | | 92818.5 | 70337.5 | | 94190 | 95967 | 88328.25 | | 0.414 |
| Lithocholic acid | 376.3 | 14 | | 440684 | 623770 | 241348 | 448497 | 438575 | | 591522 | 613289 | 634260 | 898494 | | 684391 | | 0.034 |
| Glyceraldehyde | 90.03 | 16 | | 134137451 | 149568627 | 134000070 | 43847468 | 1.2E+08 | | 61028687 | 395217386 | 86001429 | 89081725 | 98671285 | 146000102 | | 0.658 |
| m-Hydroxybenzoic acid | 138 | 16 | | 4524451.5 | 7332037.7 | | 1701793 | 4519427 | | 2458607 | 5253148.7 | 210445 | 395094 | | 2079323.8 | | 0.2 |
| Ornithine-13C | 137.1 | 18 | | 89904 | 173384 | 198804 | 299834 | 190481 | | 266153 | 138942 | 205139 | 135026 | 177764 | 184605 | | 0.893 |
| Cannabidiol | 314.2 | 14 | | 2355362.3 | 783185 | | | 1569274 | | 773234.3 | 1128193.7 | | 1140860 | | 1014096 | | 0.327 |
| 11-Hydroxytetrahydrocanna | 330.2 | 14 | | 147693.67 | 157667 | 115054 | 50933 | 117837 | | 72936.33 | 176205.5 | | 176958 | 96475 | 130643.71 | | 0.699 |
| Hydroxyglutaric acid13C | 153.1 | 12 | | 329592.5 | | | 104203 | 216898 | | 70910.67 | 287717.33 | 84270 | | | 147632.67 | | 0.532 |
| Glycine | 77.04 | 16 | | 969059.25 | 810393 | 1021475 | 356840 | 789442 | | 226956 | 389073 | | 179266 | 223654 | 254737.25 | | 0.006 |
| Glycyl-glycine | 132.1 | 18 | | 1308547 | 1488937.7 | 1626477 | 1004166 | 1357032 | | 889598.3 | 872447 | 1671246 | 847792 | 1220590 | 1100334.7 | | 0.221 |
| 8-hydroxy-2'-deoxy Guanosi | 283.1 | 18 | | 328104 | 362640.67 | 309255 | 99120 | 274780 | | 267306 | 810561.33 | 283361 | 300420 | 124096 | 357148.87 | | 0.541 |
| 5'-Methylthioadenosine | 297.1 | 12 | | 51579 | 76483 | | 126356 | 84806 | | 148197.7 | | | | 71908 | | | 0.574 |
| Inosine 2',3'-cyclic phospha | 330 | 14 | | 86845012 | 153985359 | 108102263 | 101102263 | 1.1E+08 | | 58718860 | 45392138 | 92224841 | 76370752 | 72190166 | 68979351 | | 0.015 |
| Mevalonic acid 5-phosphate | 228 | 18 | | 717803.75 | 430044.67 | 421341 | 303518 | 468177 | | 365432.3 | 618465.33 | 502558 | 431069 | 326202 | 448745.33 | | 0.83 |
| 3',5'-Cyclic Inosine monopho | 330 | 14 | | 86845012 | 153985359 | 108102263 | | 1.2E+08 | | 58718860 | 45392138 | 92224841 | 76370752 | 72190166 | 68979351 | | 0.022 |
| D-Glyceric acid | 106 | 18 | | 3096268.5 | 4352332 | 3727536 | 4900198 | 4019084 | | 2546653 | 5698507.3 | 5458965 | 3301675 | 1753035 | 3751767 | | 0.764 |
| 2-Amino-3-oxobutanoate | 117 | 16 | | 935524 | 604072.33 | | 490022 | 676539 | | 549824.3 | 1685992 | 547818 | 459627 | 683365 | 785325.27 | | 0.715 |
| Glutamic acid | 147.1 | 12 | | 5970316.5 | 6188805.3 | 6530997 | | 6230040 | | 5722491 | 4766432 | 6333652 | | | 5607525 | | 0.185 |
| 4-Methylaminobutyrate | 117.1 | 18 | | 10400593 | 13678890 | 11287937 | 7733224 | 1.1E+07 | | 6363255 | 8607741.3 | 13276841 | 8936172 | 8743777 | 9185557.2 | | 0.321 |
| Ribitol | 152.1 | 14 | | 985288.5 | 569504 | 594350 | 197602 | 586686 | | 142251 | 594437 | 559591 | 604931 | 596768 | 499595.6 | | 0.594 |
| Erythritol | 122.1 | 14 | | 1334900 | 3054234.7 | 756236 | 1511824 | 1664299 | | 4495863 | 2480118 | 3269020 | 1795571 | 4138375 | 3235789.5 | | 0.04 |
| Succinylcholine | 290.2 | 16 | | 409583.33 | 775781.5 | 601737 | | 595701 | | 268127.3 | 745956.67 | 459747 | 466458 | 329931 | 454044 | | 0.277 |
| Ethyl nitrite | 75.03 | 15 | | 1871592.8 | 1169088 | 1235187 | 713853 | 1247430 | | 1109055 | 504732 | 1221393 | 1244843 | | 1020005.8 | | 0.409 |
| 2-methyl-hexadecanedioic a | 300.2 | 17 | | 218484 | 203621 | 132689 | 61637 | 154108 | | 94677.67 | 246766 | 164077 | 209825 | 211811 | 185431.33 | | 0.446 |
| 4-Dodecylbenzenesulfonic a | 326.2 | 16 | | 1148209.8 | 834953 | | 993176 | 992113 | | 634434.3 | 370791.33 | | 385247 | 279747 | 417554.92 | | 0.001 |
| Aspartyl-Leucine | 246.1 | 14 | | 770939 | 294291.67 | 299468 | | 454900 | | 70818 | 2054826.3 | 510887 | | | 878843.78 | | 0.452 |
| AFMK | 264.1 | 19 | | 2722766 | 7320324.3 | 1809229 | 2656570 | 3627222 | | 1111438 | 5804023.3 | 3863053 | 6912415 | 3529565 | 4244098.9 | | 0.676 |
| amk | 236.1 | 12 | | 68232579 | 44656175 | 40316214 | | 5.1E+07 | | 21546886 | 223598675 | | 83388018 | | 109511193 | | 0.3 |
| Methylnoradrenaline | 183.1 | 17 | | 189741.75 | 105672 | 84976 | 81608 | 115499 | | 97991.5 | 208551 | 253894 | 209659 | 165431 | 187105.3 | | 0.065 |

#### Supplemental Figure 5

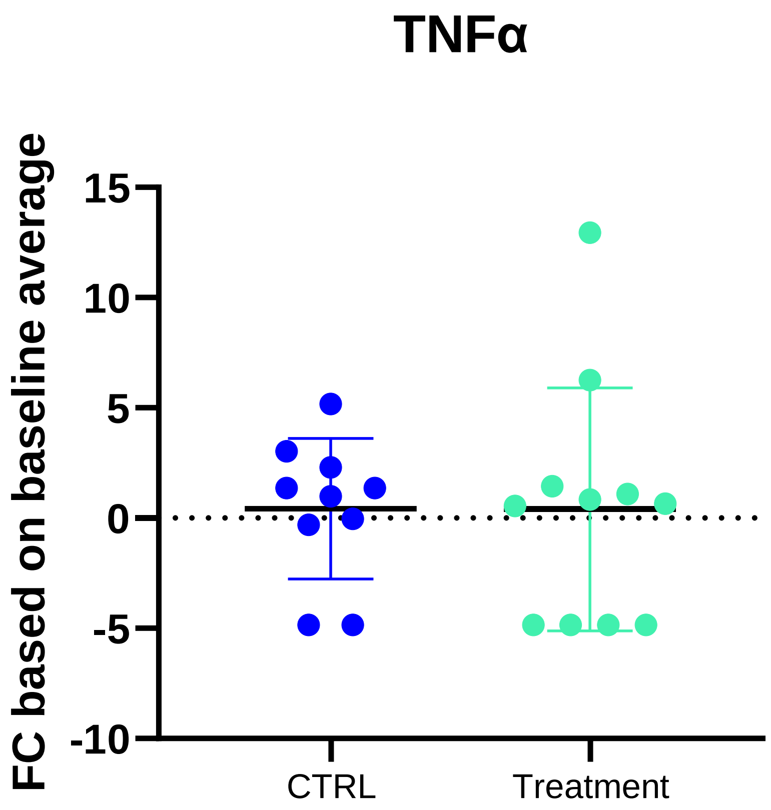


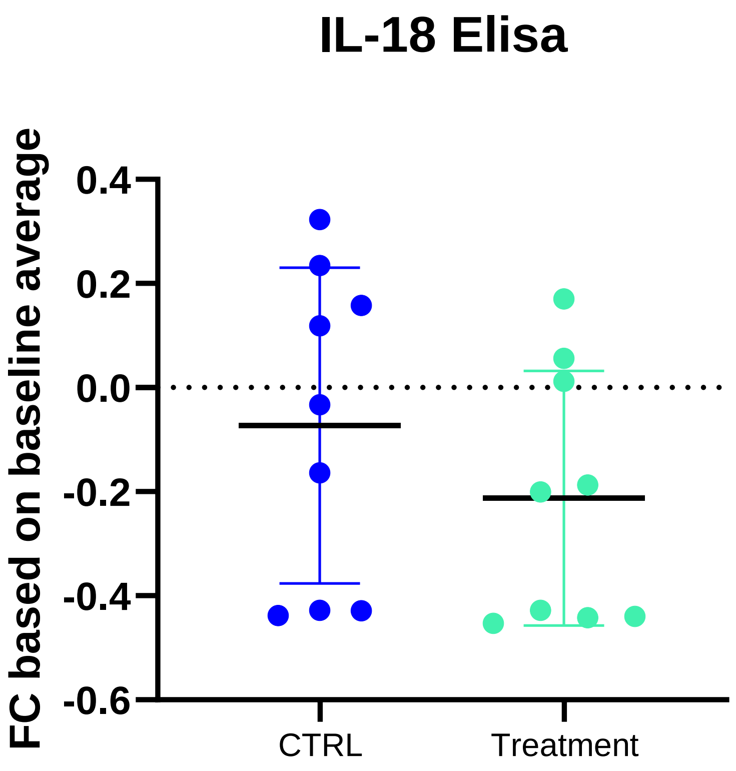


**Supplemental Figure 5**. Serum levels of Th1 cytokines TNF α and IL-18, showing treatment-induced changes.

**Supplemental Table 4**: Representativeness of Study Participants

| | |
|---|---|
| Cancer Type | Non-small cell lung cancer |
| Considerations related to: | |
| Sex | In the United States, non-small cell lung cancer (NSCLC) is more prevalent in men than women. From 2010-2017, 53% of new NSCLC cases were male. |
| Age | The median age of diagnosis for NSCLC is 71 years. For patients <65 years, the incidence per 100,000 was 13.5, and the incidence was 230.0 for patients >65 years. |
| Race/ethnicity | According to the SSER18 database, 75.2% of patients diagnosed with NSCLC were white, 12.1% were black, and 6.3% were Asian Pacific Islander. |
| Geography | From 2010-2017, there were 1.28 million new cases of NSCLC reported in the United States. |
| Other considerations | Of patients aged 55-79 diagnosed with NSCLC, 54% were diagnosed with adenocarcinoma. |
| Overall representativeness of this study | The median age for our patients was 72 years which matches the general NSCLC patient population.<br><br>50% of the patients on our study were diagnosed with adenocarcinoma.<br><br>87% of our patient population were white while 13% of our patients were black, which aligns with the NSCLC data. |
